# Leading causes of death among adult solid organ transplant recipients in the United States, 1999–2019

**DOI:** 10.1101/2024.06.07.24308602

**Authors:** Karena D Volesky-Avellaneda, Jeanny H Wang, Ruth M Pfeiffer, David Castenson, Ajay K Israni, Jonathan M Miller, Donnie Musgrove, Meredith S Shiels, Jon J Snyder, Kelly J Yu, Eric A Engels

## Abstract

Underlying medical conditions, graft failure, and immunosuppression place solid organ transplant recipients (“recipients) at heighten risk of death. We report the underlying causes of death among adult US recipients and compare their mortality to the US general population. We obtained causes of death by linking a sample of deaths from the US organ transplant registry to the National Death Index and weighted the linked deaths to represent all deaths among recipients aged ≥18 years during 1999–2019. To compare mortality to the US general population, we calculated standardized mortality ratios (SMRs). Among 496,467 recipients, 153,491 deaths occurred, of which 99,373 were NDI-linked. Leading causes of death were heart disease (16.9% of deaths), graft failure (14.9%), and cancer (14.4%). Compared to the US general population, recipients had 4.02 times the risk of death, and mortality was elevated for all 16 causes analyzed (e.g., 3.32-fold for heart disease and 2.08-fold for cancer) except dementia/Alzheimer’s disease. During 2015–2019, overall mortality was elevated 3.08-fold, with lung recipients experiencing the highest elevation (SMR=7.91), followed by heart (3.20), liver (2.86), and kidney (2.81) recipients. Although mortality improved over time, US recipients continue to face substantially elevated mortality, both overall and for common causes of death.

## INTRODUCTION

Solid organ transplantation is the treatment of choice for patients with end-stage organ disease. The United States (US) has both the highest rate and largest number of solid organ transplants performed in the world.^1^ During 1988–2023, approximately 964,000 solid organ transplants were performed in the US, with half in 2010 or later.^2^ While posttransplant survival substantially improved during the 1980s and 1990s, it has plateaued more recently, with 1-year survival remaining constant from 2012 to 2022.^3,4^ Survival also varies by the type of organ transplanted.

Acuna and colleagues conducted a study in Ontario, Canada that reported the overall distribution of causes of death (CODs) for 3068 solid organ transplant recipients (herein “recipients”) of different organ types.^5^ They found that the leading CODs were circulatory system diseases (24.0% of deaths), cancer (19.7%), and endocrine, metabolic and immune conditions (11.4%). Notably, graft failure was not listed as a possible cause of death in that study. In the US, Awan and colleagues studied 27,623 kidney recipients who died with a functioning graft during 1996–2014. Their findings revealed that cardiovascular disease followed by infections were the leading CODs; however, 56% of deaths had an unknown/missing cause.^6^

The importance of different CODs depends on the transplanted organ and likely by country, but no published studies have evaluated cause-specific mortality in the overall US recipient population. Compared to the general population, recipients are at high risk of mortality due to the medical condition necessitating the transplant, other comorbidities, failure of the transplanted organ, and immunosuppression from the medications used to prevent graft rejection. Existing studies have not compared mortality among US recipients to that of the general population.

A comprehensive assessment of CODs among recipients can inform strategies aimed at improving patient survival by identifying which causes and patient subgroups should be prioritized for interventions and further research. The US solid organ transplant registry collects COD information from transplant centers, but data are missing for a substantial proportion of deaths. For this reason, reliable information on CODs must be obtained from another source. To characterize underlying causes of mortality among adult recipients in the US, we linked a large sample of deaths listed in the US solid organ transplant registry to the National Death Index (NDI). We report the underlying CODs among adult recipients in the US, assess associations between demographic and transplant characteristics with mortality, and compare mortality among recipients to the general population.

## MATERIALS AND METHODS

Our reporting conforms to Strengthening the Reporting of Observational Studies in Epidemiology (STROBE) for cohort studies (**eTable 1** contains the STROBE checklist). This study was exempt from research ethics board review because the National Cancer Institute deemed it not human subjects research.

### Data sources, linkage, and study population

We used two data sources: the Scientific Registry of Transplant Recipients (SRTR), which has data on all solid organ transplants performed in the US since 1987,^7^ and the NDI, which includes all US death records from state vital statistic offices.^8^ To ascertain CODs, SRTR-documented deaths during 1999–2019 were linked to the NDI based on name, social security number, and date of birth. Since it was prohibitively expensive to link every death, we submitted a sample of all SRTR deaths for linkage. We sampled individuals transplanted in more recent years at a higher fraction: 40.0% of deaths among those receiving their first transplant during 1999– 2004, 75.0% for 2005–2009, and 100.0% for 2010–2019. Deaths were randomly sampled at a constant fraction within each transplant period. We followed adults (aged 18 years or older at first transplant) who lived in the continental US, Hawaii, or Puerto Rico, and received their first transplant during 1999–2019 from that transplant until death or the end of the study period on December 31, 2019 (**eTable 2**).

### Classifying causes of death

CODs provided by the NDI are based on information from death certificates. We categorized CODs into 16 groups based on the NDI-specified underlying COD, i.e., the condition/disease responsible for the chain of events leading to death (**Figure 1; eTable 3** shows how we categorized CODs into 16 groups).^9,10^ Transplant graft failure is not recorded as a COD on death certificates. To identify deaths due to graft failure, we utilized SRTR data on graft failure reported by transplant centers and assumed that deaths within 90 days of graft failure were due to graft failure. This assignment took precedence over the NDI-reported COD; however, a supplementary analysis shows how the NDI categorized recipients we classified as dying from graft failure.

**Figurer 1.**
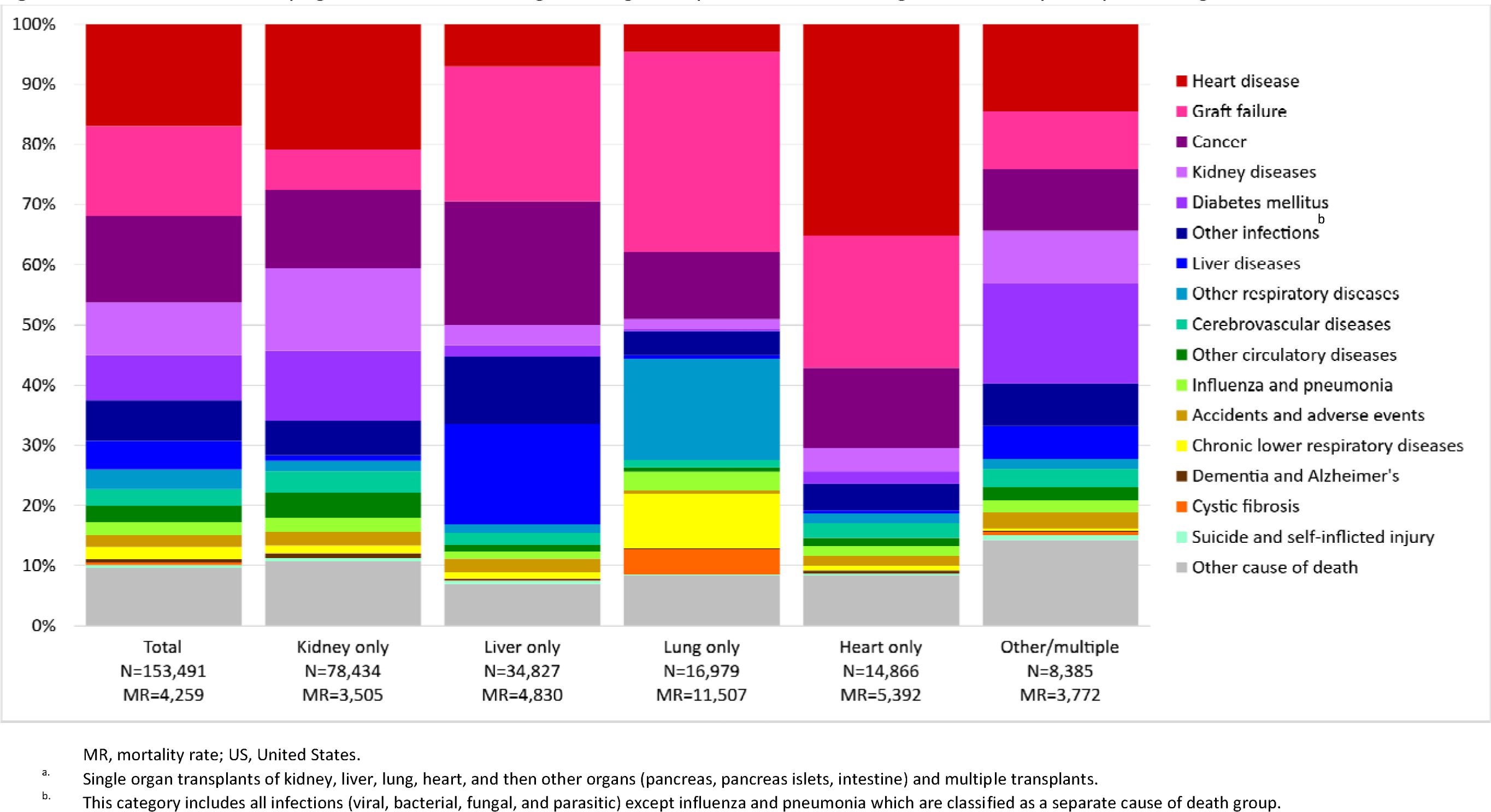
Distribution of underlying causes of death among adult organ recipients in the US during 1999–2019, by transplanted organ^a^. The 99,373 National Death Index linked deaths were weighted to represent 153,491 deaths among individuals receiving their first organ and dying during 1999–2019. N represents the number of deceased individuals who received the specified organ(s) and MR is the mortality rate per 100,000 person-years. The International Classification of Diseases 10^th^ Revision codes corresponding to the cause of death groups are reported in eTable 2.

### Statistical analysis

All analyses were performed on weighted data – 99,373 deceased recipients with NDI-linked data were weighted by the reciprocal of the final sampling fraction: 1/0.39 (for deaths among recipients transplanted during 1999–2004), 1/0.73 (2005–2009), and 1/0.98 (2010–2019). The final sampling fractions are slightly lower than we stipulated by design because a small fraction of selected SRTR deaths could not be matched (**eTable 2**). Deceased recipients who were not NDI-linked (N=54,118) were not analyzed (i.e., assigned weight of 0) because their mortality experience was captured in the weighted deaths. Recipients alive at the end of follow-up (N=342,876) were assigned a weight of 1.

The overall distribution of the 17 CODs (16 specified by NDI, plus graft failure) is reported for each of the 21 years under study and by transplanted organ for all years (1999–2019) and for the most recent year (2019). We also report how the NDI categorized recipients we classified as dying from graft failure. To assess associations of demographic and transplant characteristics with mortality, we calculated mortality rates (MRs per 100,000 person-years) and, using multivariable Poisson regression, mortality rate ratios (MRRs) for mortality overall, and for each of the 17 CODs. For the frequency, MR, and MRR analyses, we classified transplanted organs into mutually exclusive categories of four single-organ categories and an “other/multiple” group. MRRs were also calculated for graft failure by organ and for select causes depending on the organ, e.g., for kidney diseases and diabetes in kidney recipients, and heart disease in heart recipients. The Poisson regression models included the following variables captured by the SRTR: sex, age at first transplant, race and ethnicity, transplanted organ, calendar year of first transplant, and years attained since transplant (time-varying). These variables were included because they are established risk factors for many diseases.

We calculated standardized mortality ratios (SMRs), for the overall period and separately for 5-year calendar intervals. SMRs were calculated by dividing the number of observed deaths by the number of expected deaths. The expected deaths were calculated using mortality rates from the National Center for Health Statistics,^11^ stratified by sex, age, race and ethnicity, and calendar year. SMRs are also presented by transplanted organ for the study period and for 2015–2019. For the SMR analyses, we present select results for all recipients of a specified organ, regardless of whether other organs were also transplanted. For example, a person who received a lung and heart would appear in both “any lung” and “any heart” analyses. Since graft failure cannot occur in the absence of transplantation, an SMR for this outcome would not be meaningful. Instead, graft failure deaths were reclassified under the NDI-reported CODs for the SMR analyses.

Statistical calculations were performed using Stata 17 software (StataCorp) and the R *survey* package *svyglm* function.^12^ All confidence intervals (CIs) were calculated accommodating the stratified weighted sampling design. Data use agreement restrictions preclude sharing NDI data with external researchers; however, with the proper approvals, SRTR data are publicly available.^13^

## RESULTS

Among 496,367 adults who received a first transplant during 1999–2019, 62.9% were male, 60.1% were non-Hispanic White, and 57.8% were kidney recipients (**Table 1**). The median age at transplant was 54 years (interquartile range: 43–62). During 3,604,191 person-years of follow-up (median follow-up: 6.2 years), there were 153,491 deaths. A fifth (19.2%) of deaths occurred in the first year posttransplant and half (49.6%) occurred five or more years after transplantation. There were 99,373 NDI-linked deaths, reflecting the sampling design. The proportion of deaths linked to the NDI did not vary by sex or race/ethnicity but varied by age group, transplanted organ, and time since transplant. These differences reflect the sampling of deaths by calendar year of transplantation and changes over time in the composition of transplant population, as the differences were not apparent in a tabulation stratified by calendar period (**eTable 4**).

**Table 1.**
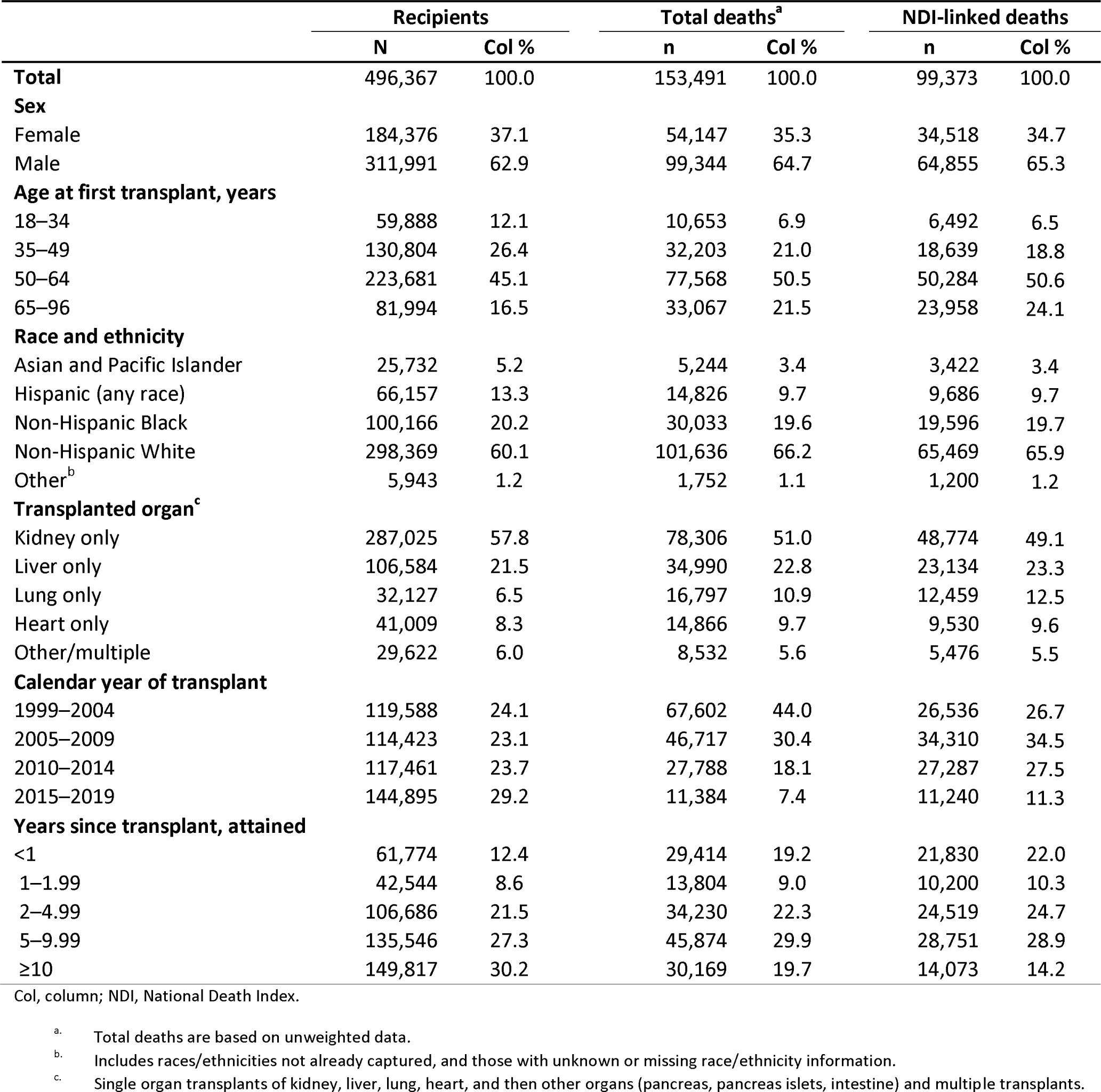
Characteristics of adult organ transplant recipients in the United States during 1999–2019.

|  | Recipients |  | Total deaths<sup>a</sup> |  | NDI-linked deaths |  |
| --- | --- | --- | --- | --- | --- | --- |
|  | N | Col % | n | Col % | n | Col % |
| **Total** | 496,367 | 100.0 | 153,491 | 100.0 | 99,373 | 100.0 |
| **Sex** |  |  |  |  |  |  |
| Female | 184,376 | 37.1 | 54,147 | 35.3 | 34,518 | 34.7 |
| Male | 311,991 | 62.9 | 99,344 | 64.7 | 64,855 | 65.3 |
| **Age at first transplant, years** |  |  |  |  |  |  |
| 18–34 | 59,888 | 12.1 | 10,653 | 6.9 | 6,492 | 6.5 |
| 35–49 | 130,804 | 26.4 | 32,203 | 21.0 | 18,639 | 18.8 |
| 50–64 | 223,681 | 45.1 | 77,568 | 50.5 | 50,284 | 50.6 |
| 65–96 | 81,994 | 16.5 | 33,067 | 21.5 | 23,958 | 24.1 |
| **Race and ethnicity** |  |  |  |  |  |  |
| Asian and Pacific Islander | 25,732 | 5.2 | 5,244 | 3.4 | 3,422 | 3.4 |
| Hispanic (any race) | 66,157 | 13.3 | 14,826 | 9.7 | 9,686 | 9.7 |
| Non-Hispanic Black | 100,166 | 20.2 | 30,033 | 19.6 | 19,596 | 19.7 |
| Non-Hispanic White | 298,369 | 60.1 | 101,636 | 66.2 | 65,469 | 65.9 |
| Other<sup>b</sup> | 5,943 | 1.2 | 1,752 | 1.1 | 1,200 | 1.2 |
| **Transplanted organ<sup>c</sup>** |  |  |  |  |  |  |
| Kidney only | 287,025 | 57.8 | 78,306 | 51.0 | 48,774 | 49.1 |
| Liver only | 106,584 | 21.5 | 34,990 | 22.8 | 23,134 | 23.3 |
| Lung only | 32,127 | 6.5 | 16,797 | 10.9 | 12,459 | 12.5 |
| Heart only | 41,009 | 8.3 | 14,866 | 9.7 | 9,530 | 9.6 |
| Other/multiple | 29,622 | 6.0 | 8,532 | 5.6 | 5,476 | 5.5 |
| **Calendar year of transplant** |  |  |  |  |  |  |
| 1999–2004 | 119,588 | 24.1 | 67,602 | 44.0 | 26,536 | 26.7 |
| 2005–2009 | 114,423 | 23.1 | 46,717 | 30.4 | 34,310 | 34.5 |
| 2010–2014 | 117,461 | 23.7 | 27,788 | 18.1 | 27,287 | 27.5 |
| 2015–2019 | 144,895 | 29.2 | 11,384 | 7.4 | 11,240 | 11.3 |
| **Years since transplant, attained** |  |  |  |  |  |  |
| <1 | 61,774 | 12.4 | 29,414 | 19.2 | 21,830 | 22.0 |
| 1–1.99 | 42,544 | 8.6 | 13,804 | 9.0 | 10,200 | 10.3 |
| 2–4.99 | 106,686 | 21.5 | 34,230 | 22.3 | 24,519 | 24.7 |
| 5–9.99 | 135,546 | 27.3 | 45,874 | 29.9 | 28,751 | 28.9 |
| ≥10 | 149,817 | 30.2 | 30,169 | 19.7 | 14,073 | 14.2 |

Col, column; NDI, National Death Index.
<sup>a</sup>. Total deaths are based on unweighted data.<sup>b</sup>. Includes races/ethnicities not already captured, and those with unknown or missing race/ethnicity information.<sup>c</sup>. Single organ transplants of kidney, liver, lung, heart, and then other organs (pancreas, pancreas islets, intestine) and multiple transplants.

Overall MRs per 100,000 person-years were 3,505 for kidney recipients, 4,830 for liver, 11,507 for lung, 5,392 for heart, and 3772 for other/multiple organ recipients. The leading CODs were heart disease, graft failure, and cancer, collectively responsible for 46.2% of recipient deaths (**Figure 1** and **eTable 5**). The leading COD was heart disease for kidney and heart recipients (20.8% and 35.1% of deaths, respectively), graft failure for liver and lung recipients (22.3% and 33.2% of deaths, respectively), and diabetes mellitus for other/multiple recipients (16.7% of deaths, driven by the inclusion of pancreas recipients; eTable 5). CODs comprising a relatively large share of deaths in recipients of a specific organ were kidney diseases (13.7% of deaths) and diabetes mellitus (11.6%) in kidney recipients, cancer (20.6%) and liver diseases (16.8%) in liver recipients, and chronic lower respiratory diseases, (9.0%) other respiratory diseases (16.9%), and cystic fibrosis (4.2%) among lung recipients.

Among all recipients, graft failure was the leading COD during 1999 through 2006, after which it was heart disease (**eTable6**). In 2011, cancer deaths overtook graft failure deaths, making cancer the second leading COD. The relative importance of cancer steadily increased year-over-year, plateauing in 2017. In 2019, the leading COD was heart disease, responsible for 17.5% of deaths, followed by cancer (16.6%) and graft failure (12.8%) (**eTable 7**). In 2019, heart disease remained the leading COD among kidney and heart recipients (22.2% and 26.3% of deaths, respectively) and graft failure the top COD among lung recipients (37.3%); however, cancer was the leading COD among liver recipients, responsible for 24.6% of deaths.

When recipients who we classified as dying from graft failure were reclassified according to the NDI-specified COD, they were often categorized under CODs related to the transplanted organ (**eTable 8**). For example, 24.4% of graft failure deaths in kidney recipients were recategorized to kidney diseases, 38.6% of graft failure deaths among liver recipients were recategorized to liver diseases, and 75.2% of graft failure deaths among heart recipients were recategorized to heart disease.

Associations with overall mortality and the three leading CODs are presented in **Table 2** (corresponding MRs reported in **eTable 9**). Compared to females, males had a 10% higher overall mortality, 27% higher heart disease mortality, and 35% higher cancer mortality. Overall mortality increased with age at first transplant, and the association with age was most pronounced for cancer and least for graft failure. Recipients who were first transplanted at the age ≥65 had more than a 4–fold higher overall mortality risk, while those transplanted at age 50–64 had 2.5-fold higher risk, compared to those transplanted at age 18–34. Non-Hispanic Black recipients experienced 20% higher overall mortality compared to non-Hispanic White recipients, whereas Hispanic and Asian and Pacific Islander recipients had 16% and 31% lower mortality, respectively. Non-Hispanic Black recipients had the highest mortality from heart disease and graft failure, whereas cancer mortality was highest among non-Hispanic White recipients.

**Table 2.**
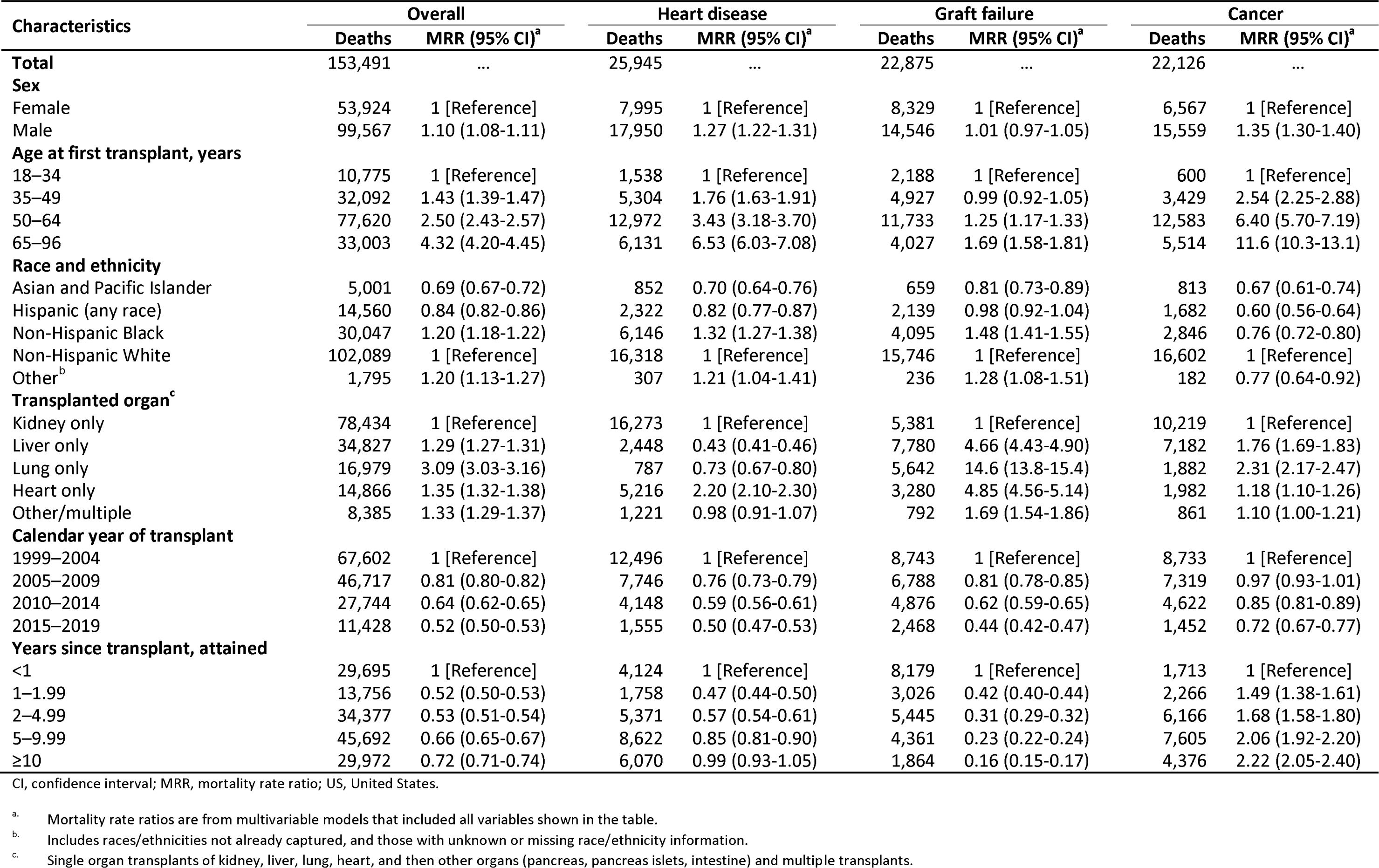
Associations of demographic and transplant characteristics with overall mortality and leading causes of death among adult organ recipients in the US during 1999–2019.

Compared to kidney recipients, overall mortality was increased by 29% for liver, 35% for heart, and 209% for lung recipients (Table 2). Heart disease mortality was more than twice as high in heart versus kidney recipients, and higher in kidney compared to liver or lung recipients. Graft failure mortality was more than 14.6-fold higher for lung recipients, and 4.66-fold higher for liver and 4.85-fold higher for heart recipients, relative to kidney recipients. Cancer mortality was most elevated in lung followed by liver and heart recipients.

Relative to transplants performed during 1999–2004, overall, heart disease, graft failure, and cancer mortality and decreased in more recent calendar periods of transplantation (Table 2). This decrease was marked for graft failure (MRR=0.44 for 2015–2019). Mortality also decreased for recipients transplanted in more recent calendar periods for all 14 remaining causes (Table 2, **eTables 10–14**). Among the three leading CODs, the decrease in mortality by calendar year of transplantation was greatest for graft failure. While there was a substantial decrease in graft failure mortality over calendar period of transplant for kidney, liver and heart recipients, there was no change for lung recipients (eTables 15, 16, 18, 19).

Overall mortality decreased by 48% after one-year posttransplant, then modestly increased in subsequent follow-up periods; however, mortality at 10 or more years posttransplant was 28% lower than in the first year (Table 2). Heart disease mortality showed a U-shaped pattern, where mortality more than halved after the first year but was the same at 10 or more years posttransplant as it was less than 1 year posttransplant. Graft failure mortality steadily declined with longer post-transplant follow-up and was 68% lower 1–1.99 years posttransplant and then 84% lower 10 or more years posttransplant than less than 1 year posttransplant. In contrast, cancer mortality steadily increased in the years following transplant and was more than double 10 or more years posttransplant versus less than 1 year posttransplant.

Among the remaining 14 causes, mortality was especially elevated for conditions related to the transplanted organ (eTables 10–14). For example, compared to kidney recipients, liver recipients experienced higher liver disease mortality (MRR=25.4), and lung recipients had higher mortality from chronic lower respiratory diseases (MRR=20.2), other respiratory diseases (MRR=27.9), and cystic fibrosis (MRR=1,103). Higher mortality in males was most pronounced for suicide and self-inflicted injury (MRR=2.72). Mortality from dementia and Alzheimer’s disease steadily increased with time since transplant, with MRRs of 25.6 at 5–9.99 years and 54.6 at 10 or more years versus less than 1 year posttransplant. The associations of demographic and transplant characteristics with select CODs, stratified by organ, are presented in **eTables 16-19**. Graft failure mortality decreased over calendar period for all organ recipients except lung recipients, among whom it remained unchanged (**eTable 18**).

Compared to the US general population, recipients had 4.02-fold higher overall mortality during 1999–2019 (**Table 3; eTable 20** presents the corresponding number of deaths and MRs). Recipients had elevated mortality from all CODs analyzed, except dementia and Alzheimer’s disease (SMR=0.82). The finding that recipients had lower dementia and Alzheimer’s disease mortality held for all organ types except kidney recipients (eTable 22). Mortality was most elevated for cystic fibrosis (SMR=389), followed by kidney diseases (19.8), other infections (11.0), other respiratory diseases (9.91), liver diseases (9.33), and diabetes (8.25). SMRs declined overall and for all CODs over calendar period; for example, the overall SMR was 5.16 in 2005–2009 and 3.08 in 2015–2019.

**Table 3.**
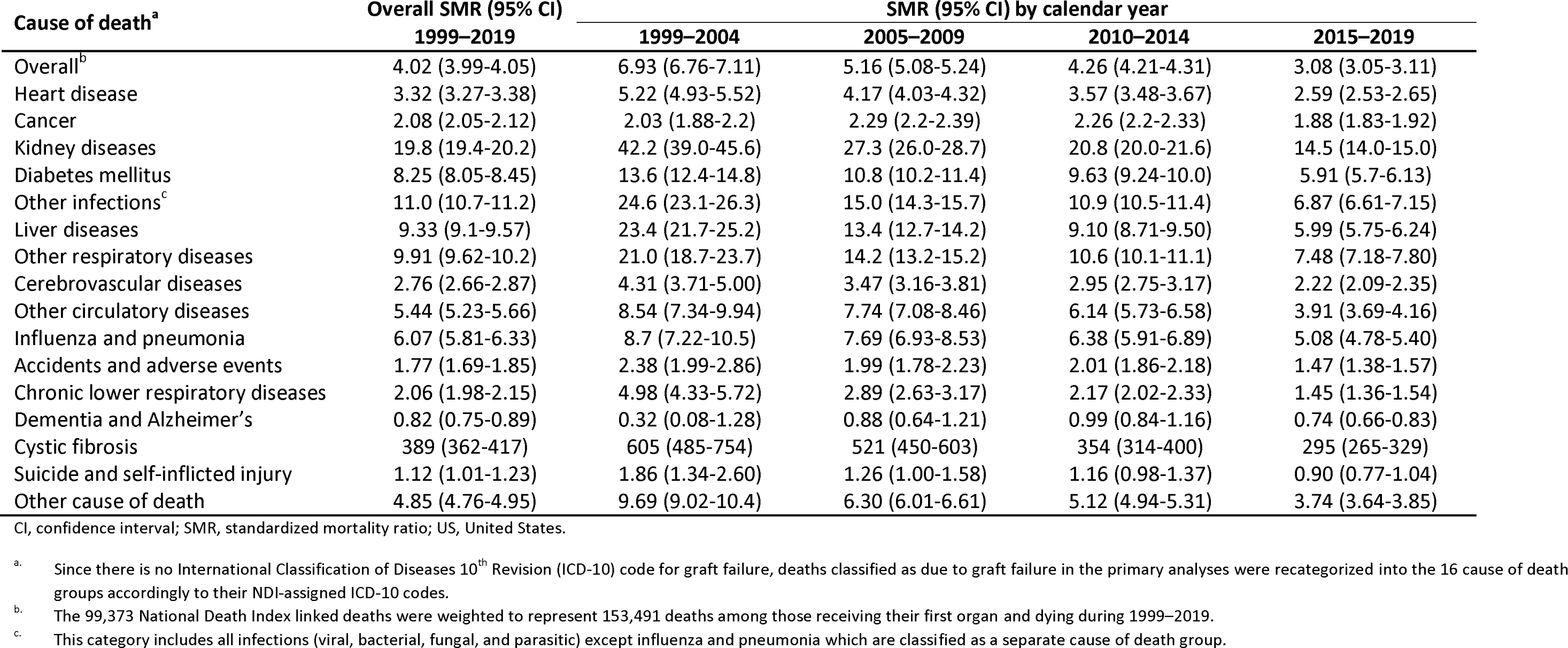
Standardized mortality ratios for causes of death among adult organ recipients in the US, overall and by calendar year.

| Cause of death<sup>a</sup> | Overall SMR (95% CI) | SMR (95% CI) by calendar year |  |  |  |
| --- | --- | --- | --- | --- | --- |
|  | 1999–2019 | 1999–2004 | 2005–2009 | 2010–2014 | 2015–2019 |
| Overall<sup>b</sup> | 4.02 (3.99-4.05) | 6.93 (6.76-7.11) | 5.16 (5.08-5.24) | 4.26 (4.21-4.31) | 3.08 (3.05-3.11) |
| Heart disease | 3.32 (3.27-3.38) | 5.22 (4.93-5.52) | 4.17 (4.03-4.32) | 3.57 (3.48-3.67) | 2.59 (2.53-2.65) |
| Cancer | 2.08 (2.05-2.12) | 2.03 (1.88-2.2) | 2.29 (2.2-2.39) | 2.26 (2.2-2.33) | 1.88 (1.83-1.92) |
| Kidney diseases | 19.8 (19.4-20.2) | 42.2 (39.0-45.6) | 27.3 (26.0-28.7) | 20.8 (20.0-21.6) | 14.5 (14.0-15.0) |
| Diabetes mellitus | 8.25 (8.05-8.45) | 13.6 (12.4-14.8) | 10.8 (10.2-11.4) | 9.63 (9.24-10.0) | 5.91 (5.7-6.13) |
| Other infections<sup>c</sup> | 11.0 (10.7-11.2) | 24.6 (23.1-26.3) | 15.0 (14.3-15.7) | 10.9 (10.5-11.4) | 6.87 (6.61-7.15) |
| Liver diseases | 9.33 (9.1-9.57) | 23.4 (21.7-25.2) | 13.4 (12.7-14.2) | 9.10 (8.71-9.50) | 5.99 (5.75-6.24) |
| Other respiratory diseases | 9.91 (9.62-10.2) | 21.0 (18.7-23.7) | 14.2 (13.2-15.2) | 10.6 (10.1-11.1) | 7.48 (7.18-7.80) |
| Cerebrovascular diseases | 2.76 (2.66-2.87) | 4.31 (3.71-5.00) | 3.47 (3.16-3.81) | 2.95 (2.75-3.17) | 2.22 (2.09-2.35) |
| Other circulatory diseases | 5.44 (5.23-5.66) | 8.54 (7.34-9.94) | 7.74 (7.08-8.46) | 6.14 (5.73-6.58) | 3.91 (3.69-4.16) |
| Influenza and pneumonia | 6.07 (5.81-6.33) | 8.7 (7.22-10.5) | 7.69 (6.93-8.53) | 6.38 (5.91-6.89) | 5.08 (4.78-5.40) |
| Accidents and adverse events | 1.77 (1.69-1.85) | 2.38 (1.99-2.86) | 1.99 (1.78-2.23) | 2.01 (1.86-2.18) | 1.47 (1.38-1.57) |
| Chronic lower respiratory diseases | 2.06 (1.98-2.15) | 4.98 (4.33-5.72) | 2.89 (2.63-3.17) | 2.17 (2.02-2.33) | 1.45 (1.36-1.54) |
| Dementia and Alzheimer's | 0.82 (0.75-0.89) | 0.32 (0.08-1.28) | 0.88 (0.64-1.21) | 0.99 (0.84-1.16) | 0.74 (0.66-0.83) |
| Cystic fibrosis | 389 (362-417) | 605 (485-754) | 521 (450-603) | 354 (314-400) | 295 (265-329) |
| Suicide and self-inflicted injury | 1.12 (1.01-1.23) | 1.86 (1.34-2.60) | 1.26 (1.00-1.58) | 1.16 (0.98-1.37) | 0.90 (0.77-1.04) |
| Other cause of death | 4.85 (4.76-4.95) | 9.69 (9.02-10.4) | 6.30 (6.01-6.61) | 5.12 (4.94-5.31) | 3.74 (3.64-3.85) |

CI, confidence interval; SMR, standardized mortality ratio; US, United States.
<sup>a</sup> Since there is no International Classification of Diseases 10<sup>th</sup> Revision (ICD-10) code for graft failure, deaths classified as due to graft failure in the primary analyses were recategorized into the 16 cause of death groups accordingly to their NDI-assigned ICD-10 codes.
<sup>b</sup> The 99,373 National Death Index linked deaths were weighted to represent 153,491 deaths among those receiving their first organ and dying during 1999–2019.
<sup>c</sup> This category includes all infections (viral, bacterial, fungal, and parasitic) except influenza and pneumonia which are classified as a separate cause of death group.

For each COD, the SMRs in 2015–2019 were almost all lower than they were for 2010–2014. Recipients were at elevated risk of death from suicide only in 1999–2004. Declines in SMRs over calendar period were marked for kidney diseases, other infections, liver diseases, and chronic respiratory diseases. The elevation in chronic lower respiratory disease mortality (SMR=2.06) was entirely driven by lung recipients (SMR=25.5) because mortality for this outcome was decreased in kidney, liver, and heart recipients relative to the general population.

Overall SMRs greatly varied by organ transplanted, ranging from a 3.46-fold elevation (2.81-fold in 2015–2019) for kidney recipients to a 10.5-fold elevation (7.91-fold in 2015–2019) for lung recipients (**Table 4, eTable 21**). Among kidney recipients, mortality was most increased for kidney diseases (SMR=24.3), followed by diabetes (SMR=11.3). For liver recipients, mortality was especially elevated for liver diseases (SMR=32.9) and other infections (SMR=25.7). Among lung recipients, the highest SMRs were for cystic fibrosis (SMR=7420), other respiratory diseases (SMR=149), and influenza/pneumonia (SMR=32.4). For heart recipients, mortality from kidney diseases (SMR=9.9) and heart disease (SMR=8.77) were notably increased.

**Table 4.** Standardized mortality ratios for causes of death among adult organ recipients in the US during 1999–2019 and 2015–2019, by transplanted organ SMR (95% CI), by transplanted organ.

| Cause of death | SMR (95% CI), by transplanted organ<sup>a</sup> |  |  |  |  |  |  |  |
| --- | --- | --- | --- | --- | --- | --- | --- | --- |
|  | Kidney |  | Liver |  | Lung |  | Heart |  |
|  | 1999–2019 | 2015–2019 | 1999–2019 | 2015–2019 | 1999–2019 | 2015–2019 | 1999–2019 | 2015–2019 |
| Overall | 3.46<br>(3.43-3.49) | 2.81<br>(2.77-2.85) | 4.38<br>(4.32-4.45) | 2.86<br>(2.8-2.92) | 10.5<br>(10.3-10.7) | 7.91<br>(7.70-8.12) | 4.16<br>(4.07-4.25) | 3.20<br>(3.10-3.30) |
| Heart disease | 3.14<br>(3.08-3.2) | 2.56<br>(2.48-2.63) | 1.47<br>(1.4-1.55) | 1.30<br>(1.22-1.39) | 2.98<br>(2.77-3.21) | 2.48<br>(2.24-2.74) | 8.77<br>(8.51-9.04) | 6.00<br>(5.72-6.28) |
| Cancer | 1.58<br>(1.54-1.62) | 1.49<br>(1.44-1.54) | 3.19<br>(3.1-3.28) | 2.4<br>(2.39-2.59) | 4.02<br>(3.82-4.24) | 4.04<br>(3.77-4.32) | 1.83<br>(1.73-1.94) | 1.73<br>(1.59-1.88) |
| Kidney diseases | 24.3<br>(23.7-24.9) | 17.5<br>(16.9-18.2) | 9.75<br>(9.09-10.5) | 7.74<br>(7.01-8.54) | 13.4<br>(11.9-15.2) | 12.4<br>(10.6-14.6) | 9.89<br>(8.93-11.0) | 8.46<br>(7.31-9.79) |
| Diabetes mellitus | 11.3<br>(11.0-11.5) | 8.07<br>(7.76-8.39) | 2.43<br>(2.21-2.67) | 1.99<br>(1.74-2.28) | 1.32<br>(1-1.74) | 1.35<br>(0.97-1.89) | 2.59<br>(2.25-2.98) | 2.10<br>(1.71-2.58) |
| Other infections<sup>b</sup> | 6.72<br>(6.49-6.96) | 5.83<br>(5.53-6.15) | 25.7<br>(24.8-26.5) | 9.39<br>(8.76-10.1) | 19.1<br>(17.6-20.7) | 14.07<br>(12.5-15.9) | 7.2<br>(6.57-7.9) | 5.47<br>(4.75-6.3) |
| Liver diseases | 1.98<br>(1.85-2.12) | 1.65<br>(1.5-1.83) | 32.9<br>(32.1-33.8) | 19.7<br>(18.8-20.5) | 2.54<br>(2.03-3.17) | 1.84<br>(1.29-2.62) | 0.89<br>(0.69-1.16) | 0.78<br>(0.54-1.14) |
| Other respiratory diseases | 3.23<br>(3.02-3.46) | 2.75<br>(2.51-3.02) | 3.58<br>(3.22-3.97) | 2.71<br>(2.34-3.15) | 149<br>(145-155) | 101<br>(96.2-106) | 4.03<br>(3.47-4.66) | 3.31<br>(2.70-4.07) |
| Cerebrovascular diseases | 2.81<br>(2.67-2.94) | 2.33<br>(2.18-2.5) | 2.38<br>(2.17-2.6) | 1.75<br>(1.53-2.00) | 3.85<br>(3.29-4.5) | 2.91<br>(2.33-3.63) | 2.67<br>(2.34-3.05) | 2.01<br>(1.64-2.47) |
| Other circulatory diseases | 6.55<br>(6.27-6.85) | 4.79<br>(4.48-5.13) | 3.05<br>(2.73-3.4) | 2.14<br>(1.81-2.52) | 4.25<br>(3.47-5.21) | 3.43<br>(2.6-4.54) | 3.34<br>(2.83-3.94) | 2.30<br>(1.78-2.97) |
| Influenza and pneumonia | 5.38<br>(5.09-5.7) | 4.67<br>(4.31-5.06) | 3.81<br>(3.4-4.27) | 3.36<br>(2.87-3.93) | 32.4<br>(29.7-35.4) | 22.7<br>(20.0-25.8) | 4.64<br>(3.95-5.46) | 4.4<br>(3.51-5.51) |
| Accidents and adverse events | 1.61<br>(1.52-1.71) | 1.32<br>(1.22-1.44) | 2.33<br>(2.14-2.53) | 1.95<br>(1.73-2.2) | 2.00<br>(1.63-2.45) | 1.91<br>(1.47-2.48) | 1.56<br>(1.33-1.84) | 1.32<br>(1.05-1.66) |
| Chronic lower respiratory diseases | 0.83<br>(0.76-0.9) | 0.73<br>(0.65-0.82) | 0.77<br>(0.67-0.88) | 0.66<br>(0.54-0.8) | 25.5<br>(24.2-26.8) | 14.1<br>(13.1-15.3) | 0.64<br>(0.51-0.8) | 0.73<br>(0.55-0.98) |
| Dementia and Alzheimer's | 0.90<br>(0.81-1.00) | 0.82<br>(0.72-0.93) | 0.51<br>(0.39-0.67) | 0.47<br>(0.34-0.65) | 0.62<br>(0.38-1.02) | 0.8<br>(0.46-1.39) | 0.77<br>(0.56-1.07) | 0.64<br>(0.41-0.99) |
| Cystic fibrosis | 8.97<br>(4.86-16.6) | 5.01<br>(1.86-13.5) | 39.3<br>(24.0-64.3) | 44.8<br>(23.9-84.2) | 7420<br>(6921-7955) | 5473<br>(4901-6111) | 18.4<br>(5.43-62.1) | <0.01<sup>c</sup> |
| Suicide and self-inflicted injury | 1.07<br>(0.94-1.21) | 0.81<br>(0.67-0.99) | 1.41<br>(1.20-1.67) | 1.23<br>(0.97-1.57) | 1.15<br>(0.76-1.76) | 1.05<br>(0.55-2.00) | 0.68<br>(0.46-1.00) | 0.51<br>(0.26-1.01) |
| Other cause of death | 4.32<br>(4.21-4.44) | 3.39<br>(3.26-3.53) | 4.35<br>(4.16-4.54) | 3.09<br>(2.9-3.3) | 13.9<br>(13.2-14.7) | 10.5<br>(9.78-11.3) | 5.26<br>(4.95-5.58) | 4.25<br>(3.91-4.62) |

CI, confidence interval; SMR, standardized mortality ratio; US, United States.
<sup>a</sup>. All recipients of a specified organ, regardless of whether other organs were also transplanted.<sup>b</sup>. This category includes all infections (viral, bacterial, fungal, and parasitic) except influenza and pneumonia which are classified as a separate cause of death group.<sup>c</sup>. Since there were no deaths due to cystic fibrosis among heart recipients during 2015–2019, the SMR estimate was extremely small.

## DISCUSSION

This study provides the first broad and systematic overview of CODs among adult recipients in the US over more than two decades. We demonstrate that heart disease, graft failure, and cancer were the most common causes of mortality among US recipients during 1999–2019. The importance of these and other CODs varied considerably depending on the transplanted organ.

Reassuringly, recipients receiving their first transplant in more recent years had lower mortality for all 17 CODs analyzed. These decreases in mortality reflect changes in medical management over time, including better organ matching, transplant techniques, immunosuppressive regimens, infection prophylaxis and treatment, and surveillance and long-term care. Despite improvements in transplant care over time, mortality in recipients remained 3.08-fold higher during 2015–2019 than in the general US population. Of note, the mortality patterns we described for calendar year of transplant, years attained since transplant, and calendar year of death are inextricably linked. For example, deaths during 1999–2004 occurred among individuals transplanted at most five years earlier, while deaths during 2015–2019 include those that occurred in recipients first transplanted as many as two decades earlier. Readers should keep this in mind when interpreting trends over time and the distribution of CODs at the start of the study period.

Aging of the recipient population also contributed to the observed mortality patterns over time. In this study, the median age at first transplantation was 49 years in 1999 and 56 years in 2019. The older age of recipients, coupled with decreasing graft failure mortality, likely explains why heart disease emerged as the leading COD in the mid-2000s. In the general population, heart disease has been the leading COD for decades, and in 2019 accounted for 23.1% of deaths.^14^ Here, heart disease compromised 17.5% of deaths in 2019. Heart disease was the leading COD in kidney and heart recipients. This finding is consistent with prior reports of kidney recipients in the US, Australia and New Zealand, and Spain.^6,15,16^ Heart disease risk factors are especially common in kidney and heart recipients. For example, in 2019, 38% and 21% of US kidney recipients had a primary diagnosis of diabetes or hypertension, respectively.^17^ Kidney recipients also experience chronic inflammation which can damage blood vessels thereby increasing the risk of cardiovascular complications. In addition, some immunosuppressive therapies administered to prevent graft rejection have side effects including hypertension, hypercholesterolemia, and diabetes. In heart recipients, cardiac allograft vasculopathy occurs in about half of heart recipients within 10 years of transplant and can lead to death.^18^

Graft failure was the second leading COD across organ types during 1999–2019, but the leading COD for liver and lung recipients. Mortality from graft failure halved after 1 year posttransplant and steadily decreased with longer time since transplant. Improvements in graft survival over calendar time are well documented,^19,20^ with improved surgical techniques, better organ matching, and superior immunosuppressive therapies all contributing. Among the leading CODs, the decrease in mortality across calendar periods was most pronounced for graft failure. In Copenhagen, Denmark, Søborg et al. reported that the decrease they observed in graft failure deaths during 2010–2020 was driven by a decrease in graft failure in lung recipients. In contrast, we found that the decrease in graft failure mortality was limited to kidney, liver and heart recipients.

Cancer became an increasingly common COD over time and was the second leading COD during 2011–2019. Due to immunosuppression, recipients have elevated incidence of posttransplant cancers and poor survival after a cancer diagnosis, and we recently reported on cancer mortality in US recipients in detail.^21^ Here, we describe that cancer mortality steadily increased with time since transplant, likely reflecting the effects of prolonged immunosuppression. In addition, hepatocellular carcinoma is a common indication for liver transplantation in the US,^22^ and the risk of recurrence after transplantation is high, contributing to the high cancer mortality in liver recipients. Finally, kidney and lung recipients have a high risk of developing cancer in the damaged native organs that may be left in place during transplantation.

By comparing mortality risks in the transplant population to the general US population, we found that recipients were at higher mortality risk for all but one of the 16 COD groups, reflecting the high prevalence of medical comorbidity in the recipient population and complications of transplantation, including those related to immunosuppression. Dementia and Alzheimer’s disease was the only COD where the SMR was reduced. Patients referred to a transplant center undergo a thorough medical and psychosocial evaluation, and patients with advanced dementia or Alzheimer’s disease would not be deemed candidates for organ transplant. Dementia and Alzehimer’s disease was the only COD where the risk of death greatly increased with time since transplant. This may be because of this screening process whereby individuals with this disease are initially screened out, but over time new cases of the disease could become manifest and lead to death.

Compared to recipients of other organs, lung recipients had by far the highest overall mortality and mortality due to graft failure. Examining US transplants performed 1990–2018, Graham and colleagues estimated that mean survival posttransplant was only 9.3 years for lung recipients, compared with 22.8 and 20.9 years for kidney and liver recipients, respectively.^23^ In particular bronchiolitis obliterans (a form of chronic allograft rejection) is common after lung transplantation.^24^ Exposure of the transplanted lung to the environment coupled with an impaired cough reflex leaves lung recipients at high risk of infection. Compared to the general population, we found that lung recipients had vastly elevated mortality from infections, respiratory diseases, and cystic fibrosis. In 2019, cystic fibrosis was the primary diagnoses for ∼10% of lung transplantations in the US.^24^ The extremely elevated SMR for this disease thus arises because cystic fibrosis is an indication for lung transplantation and due to its extreme rarity in the population (i.e., estimated prevalence of 40,000 people in 2020).^25^

There are several strengths of our study to highlight. Transplant centers are required to provide annual follow-up reports on recipient vital status, and the SRTR identifies additional deaths through linkage to the National Technical Information Service’s Death Master File and the Centers for Medicare and Medicaid Services. Our study thus captures virtually all US recipient deaths. Our linkage to the NDI yielded uniquely valuable COD data, as a large fraction of recipients are missing center-provided CODs in the SRTR (43.8% of recipients in this study population). Further, by linking to the NDI and accounting for graft failure, we provided a comprehensive portrait of mortality in this population, including less common CODs such as dementia and Alzheimer’s disease and cystic fibrosis. We were also able to compare CODs among recipients of different organ types.

Because this study was a broad overview, we could not go into detail for specific CODs. Since we restricted the study population to first transplants during 1999–2019, these results may not be generalizable to recipients transplanted before 1999. Graft failure is not listed in the NDI, so we assumed that deaths within 90 days of SRTR-documented graft failure were graft failure deaths. Leading CODs among children and adults differ. We therefore did not combine these groups and instead focused this study on adults. Some risk factors for mortality, such as lifestyle factors and immunosuppressive regimens, were not captured or analyzed. Finally, our findings cannot be extrapolated to 2020 and beyond because of the emergence of COVID-19, which was responsible for ∼20% of recipient deaths in 2020–2021.^26,27^

In conclusion, mortality among US recipients has improved in recent calendar periods, but recipients remain at elevated risk of death compared to the general population. Based on our results, we highlight three areas for further research. First, relative to non-Hispanic White recipients, non-Hispanic Black recipients had worse overall mortality, and mortality was most increased in non-Hispanic Black recipients for graft failure, heart and kidney diseases, other circulatory diseases, and infections other than influenza and pneumonia. Second, graft failure remains an especially common COD in lung recipients (37.3% of deaths in this group in 2019). Third, lung recipients had markedly elevated mortality for several outcomes. Targeted studies of these populations and outcomes could help lead to improved recipient survival.

## Supporting information

Supplementary Content

## Acknowledgements / Funding

Intramural Research Program of the National Cancer Institute.

This work was conducted partially under the auspices of the Hennepin Healthcare Research Institute (HHRI), contractor for the Scientific Registry of Transplant Recipients (SRTR), under contract no. 75R60220C00011 (US Department of Health and Human Services, Health Resources and Services Administration, Healthcare Systems Bureau, Division of Transplantation). The US Government (and others acting on its behalf) retains a paid-up, nonexclusive, irrevocable, worldwide license for all works produced under the SRTR contract, and to reproduce them, prepare derivative works, distribute copies to the public, and perform publicly and display publicly, by or on behalf of the Government. The data reported here have been supplied by HHRI as the contractor for SRTR. The interpretation and reporting of these data are the responsibility of the author(s) and in no way should be seen as an official policy of or interpretation by SRTR or the US Government.

## Data availability statement

Data use agreement restrictions preclude sharing study data with external researchers.

## Supporting information statement

Additional supporting information may be found online in the Supporting Information section.

## Acronyms and abbreviations

CI: confidence interval
COD: cause of death
ICD-10: International Classification of Diseases 10^th^ Revision
MR: mortality rate
MRR: mortality rate ratio
NDI: National Death Index
SMR: standardized mortality ratio
SRTR: Scientific Registry of Transplant Recipients
US: United States

