## Supplementary Content for "Leading causes of death among adult solid organ transplant recipients in the United States, 1999–2019"

### eTable 1. STROBE Statement – Checklist for cohort studies

|  | Item no | Recommendation | Page no |
| --- | --- | --- | --- |
| **Title and abstract** | 1 | (*a*) Indicate the study’s design with a commonly used term in the title or the abstract | 4 |
|  |  | (*b*) Provide in the abstract an informative and balanced summary of what was done and what was found |  |
| Introduction | | | |
| Background/rationale | 2 | Explain the scientific background and rationale for the investigation being reported | 5 |
| Objectives | 3 | State specific objectives, including any prespecified hypotheses | 5 |
| Methods | | | |
| Study design | 4 | Present key elements of study design early in the paper | 6 |
| Setting | 5 | Describe the setting, locations, and relevant dates, including periods of recruitment, exposure, follow–up, and data collection | 6 |
| Participants | 6 | (*a*) Give the eligibility criteria, and the sources and methods of selection of participants. Describe methods of follow–up | 6  eTable 2 |
|  |  | (*b*) For matched studies, give matching criteria and number of exposed and unexposed | NA |
| Variables | 7 | Clearly define all outcomes, exposures, predictors, potential confounders, and effect modifiers. Give diagnostic criteria, if applicable | 6 & 7 |
| Data sources/ measurement | 8* | For each variable of interest, give sources of data and details of methods of assessment (measurement). Describe comparability of assessment methods if there is more than one group | 7 |
| Bias | 9 | Describe any efforts to address potential sources of bias | 7 |
| Study size | 10 | Explain how the study size was arrived at | eTable 2 |
| Quantitative variables | 11 | Explain how quantitative variables were handled in the analyses. If applicable, describe which groupings were chosen and why | 7 |
| Statistical methods | 12 | (*a*) Describe all statistical methods, including those used to control for confounding | 7 |
|  |  | (*b*) Describe any methods used to examine subgroups and interactions |  |
|  |  | (*c*) Explain how missing data were addressed |  |
|  |  | (*d*) If applicable, explain how loss to follow–up was addressed |  |
|  |  | (*e*) Describe any sensitivity analyses |  |

STROBE, Strengthening the Reporting of Observational Studies in Epidemiology. **eTable 1. STROBE Statement – Checklist for cohort studies**

|  | **Item no** | **Recommendation** | **Page no** |
| --- | --- | --- | --- |
| **Results** | | | |
| Participants | 13* | (a) Report numbers of individuals at each stage of study—eg numbers potentially eligible, examined for eligibility, confirmed eligible, included in the study, completing follow–up, and analysed | eTable 2 |
|  |  | (b) Give reasons for non–participation at each stage |  |
|  |  | (c) Consider use of a flow diagram |  |
| Descriptive data | 14* | (a) Give characteristics of study participants (eg demographic, clinical, social) and information on exposures and potential confounders | 8  Table 1 |
|  |  | (b) Indicate number of participants with missing data for each variable of interest |  |
|  |  | (c) Summarise follow–up time (eg, average and total amount) |  |
| Outcome data | 15* | Report numbers of outcome events or summary measures over time | Table 2, 3, 4  eTable 7 |
| Main results | 16 | (*a*) Give unadjusted estimates and, if applicable, confounder–adjusted estimates and their precision (eg, 95% confidence interval). Make clear which confounders were adjusted for and why they were included | Table 2, 3, 4  eTables 9–19 |
|  |  | (*b*) Report category boundaries when continuous variables were categorized |  |
|  |  | (*c*) If relevant, consider translating estimates of relative risk into absolute risk for a meaningful time period |  |
| Other analyses | 17 | Report other analyses done—eg analyses of subgroups and interactions, and sensitivity analyses | eTables 9–25 |
| Discussion | | | |
| Key results | 18 | Summarise key results with 1 [Reference] to study objectives | 12 |
| Limitations | 19 | Discuss limitations of the study, taking into account sources of potential bias or imprecision. Discuss both direction and magnitude of any potential bias | 12–15 |
| Interpretation | 20 | Give a cautious overall interpretation of results considering objectives, limitations, multiplicity of analyses, results from similar studies, and other relevant evidence | 12–15 |
| Generalis-ability | 21 | Discuss the generalisability (external validity) of the study results | 15 |
| Other information | | | |
| Funding | 22 | Give the source of funding and the role of the funders for the present study and, if applicable, for the original study on which the present article is based | 3 |

STROBE, Strengthening the Reporting of Observational Studies in Epidemiology.

### eTable 2. Study population selection

|  | **People receiving a first transplant, N** | **Deaths, N** | **Deaths linked to NDI data, N** |
| --- | --- | --- | --- |
| **SRTR data used for selecting deaths for NDI linkage, transplants during 1987–2019** | 711,276 | 291,385 | 138,664 |
| Transplants and deaths included in the October 2020 SRTR Standard Analytic File,  with first transplant during 1987-2019 | 711,276 | 289,114^b^ | 138,664 |
| **First transplant during 1999–2019^a^** | 533,119 | 160,695 | 103,910 |
| **Exclusion of** |  |  |  |
| Duplicate records | 533,116 | 160,692 | 103,908 |
| Records with missing or conflicting data on age or sex | 533,059 | 160,679 | 103,904 |
| Those with unknown or conflicting information about region of residence | 532,453 | 160,432 | 103,814 |
| Those residing outside of continental US, Hawaii, or Puerto Rico | 530,608 | 160,120 | 103,710 |
| Those who died on the same day as transplant | 529,158 | 158,779 | 102,723 |
| Those aged <18 years at transplant | **496,367** | **153,491** | **99,373** |

NDI, National Death Index; SRTR, Scientific Registry of Transplant Recipients.

a. From a larger linkage of SRTR and NDI data involving transplants and deaths during 1987–2018, we applied an additional exclusion criterion of restricting to transplants and deaths from 1999–2019 because most recent data are most relevant for informing interventions and research priorities.
b. Excluded 2,271 deaths that were restricted because they were identified only in the social security file and not released in the SRTR Standard Analytic File.

### eTable 3. Classification of causes of death

| **Cause of death** | **ICD–10 (1999 onwards)^a,b^** |
| --- | --- |
| **Heart disease** |  |
| Acute rheumatic fever, chronic rhematic fever | I00–I09 |
| Hypertensive heart disease; hypertensive heart and renal disease | I11; I13 |
| Ischaemic heart diseases, pulmonary heart disease and diseases of pulmonary circulation, other forms of heart disease | I20–I51 |
| **Neoplasms (cancer)** |  |
| Malignant | C00–C97 |
| In situ neoplasms; benign neoplasms; neoplasms of uncertain or unknown behaviour | D00–D09; D10–D36; D37–D48 |
| **Kidney diseases** |  |
| Nephritis, nephrotic syndrome and nephrosis | N00–N07, N17–N19, N25–N27 |
| Renal tubulo–interstitial diseases | N10–N16 |
| Urolithiasis; Other disorders of kidney and ureter | N20–N23; N28 |
| **Diabetes mellitus** | E10–E14 |
| **Other infections** |  |
| Intestinal infectious diseases | A00–A09 |
| Tuberculosis | A15–A19 |
| Certain zoonotic bacterial diseases | A20–A28 |
| Other bacterial diseases | A30–A49 |
| Infections with a predominantly sexual mode of transmission | A50–A64 |
| Viral hepatitis | B15–B19 |
| Human immunodeficiency virus | B20–B24 |
| Other viral diseases | B25–B34 |
| Mycoses; protozoal diseases; helminthiases | B35–B49; B50–B64; B65–B83 |
| Pediculosis, acariasis and other infestations; | B85–B89 |
| Sequelae of infectious and parasitic diseases | B90–B98 |
| Bacterial, viral and other infectious agents | B95–B98 |
| Other and unspecified infectious diseases | B99 |
| **Liver diseases** |  |
| Chronic liver disease and cirrhosis | K70, K73, K74 |
| Toxic liver disease; Hepatic failure, not elsewhere classified | K71; K72 |
| Other inflammatory liver diseases; other diseases of liver | K75; K76 |
| **Other respiratory system diseases** |  |
| Acute upper respiratory infections | J00–J06 |
| Other acute lower respiratory infections | J20–J22 |
| Other diseases of upper respiratory tract | J30–J39 |
| Lung diseases due to external agents | J60–J70 |
| Other respiratory diseases principally affecting the interstitium | J80–J84 |
| Suppurative and necrotic conditions of lower respiratory tract | J85–J86 |
| Other diseases of pleura; other diseases of the respiratory system | J90–J94; J95–J99 |
| **Cerebrovascular diseases** | I60–I69 |
| **Other circulatory system diseases** |  |
| Hypertension without heart disease | I10, I12, I15 |
| Diseases of arteries, arterioles and capillaries | I70–I78 |
| Diseases of veins, lymphatic vessels and lymph nodes, not elsewhere classified | I80–I89 |
| Other and unspecified disorders of the circulatory system | I95–I99 |
| **Influenza and pneumonia** | J09–J18 |
| **Accidents and adverse effects** |  |
| Accidents | V01–X59 |
| Assault by drugs, medicaments and biological substances; assault by corrosive substance | Y85; Y86 |
| **Chronic lower respiratory diseases** | J40–J47 |
| **Dementia and Alzheimer’s disease** |  |
| Dementia in Alzheimer disease; Vascular dementia; Dementia in other diseases classified elsewhere; Unspecified dementia | F00– F03 |
| Alzheimer’s disease | G30 |
| **Cystic fibrosis** | E84 |
| **Suicide and self–inflicted injury** | U03, X60–X84, Y87.0 |
| All remaining ICD–10 codes | … |

a. ICD-10 International Classification of Diseases 10^th^ Revision
b. Since using the Surveillance, Epidemiology, and End Results Program (SEER) Cause of Death Recode 1969+^1^ categories left ~30% of deaths in the ‘all other’ category, we re-classified certain CODs under existing SEER categories (e.g., dementia was combined with Alzheimer’s disease) and created addition COD groups: other respiratory system diseases, other circulatory system diseases, and cystic fibrosis.

### eTable 4. Proportion of deaths among adult organ recipients in the US submitted for NDI record linkage, by demographic and transplant characteristics, 1999–2019

|  |  | | | | |  | **SRTR documented deaths by calendar period of first transplant** | | | | | | | | | | | | | | | | |
| --- | --- | --- | --- | --- | --- | --- | --- | --- | --- | --- | --- | --- | --- | --- | --- | --- | --- | --- | --- | --- | --- | --- | --- |
|  | **All deaths in the SRTR**  **153,491 deaths**  **NDI-linked** | | | | |  | **1999–2004**  **67,602 deaths**  **NDI-linked** | | | | |  | **2005–2009**  **46,717 deaths**  **NDI-linked** | | | | | **2010–2019**  **39,172 deaths**  **NDI-linked** | | | | | |
| **Characteristics** | **No** | |  | **Yes** | |  | **No** | |  | **Yes** | |  | **No** | |  | **Yes** | |  | **No** | |  | **Yes** | |
|  | **N** | **Col %** |  | **N** | **Col %** |  | **N** | **Col %** |  | **N** | **Col %** |  | **N** | **Col %** |  | **N** | **Col %** |  | **N** | **Col %** |  | **N** | **Col %** |
| **Total** | 54,118 |  |  | 99,373 |  |  | 41,066 |  |  | 26,536 |  |  | 12,407 |  |  | 34,310 |  |  | 645 |  |  | 38,527 |  |
| **Sex** |  |  |  |  |  |  |  |  |  |  |  |  |  |  |  |  |  |  |  |  |  |  |  |
| Female | 19,629 | 36.3 |  | 34,518 | 34.7 |  | 15,219 | 37.1 |  | 9,672 | 36.4 |  | 4,212 | 33.9 |  | 11,662 | 34.0 |  | 198 | 30.7 |  | 13,184 | 34.2 |
| Male | 34,489 | 63.7 |  | 64,855 | 65.3 |  | 25,847 | 62.9 |  | 16,864 | 63.6 |  | 8,195 | 66.1 |  | 22,648 | 66.0 |  | 447 | 69.3 |  | 25,343 | 65.8 |
| **Age at first transplant, years** | | |  |  |  |  |  |  |  |  |  |  |  |  |  |  |  |  |  |  |  |  |  |
| 18–34 | 4,161 | 7.7 |  | 6,492 | 6.5 |  | 3,313 | 8.1 |  | 2,257 | 8.5 |  | 804 | 6.5 |  | 2,086 | 6.1 |  | 44 | 6.8 |  | 2,149 | 5.6 |
| 35–49 | 13,564 | 25.1 |  | 18,639 | 18.8 |  | 11,279 | 27.5 |  | 7,177 | 27.0 |  | 2,187 | 17.6 |  | 6,248 | 18.2 |  | 98 | 15.2 |  | 5,214 | 13.5 |
| 50–64 | 27,284 | 50.4 |  | 50,284 | 50.6 |  | 20,431 | 49.8 |  | 13,230 | 49.9 |  | 6,559 | 52.9 |  | 18,099 | 52.8 |  | 294 | 45.6 |  | 18,955 | 49.2 |
| 65–96 | 9,109 | 16.8 |  | 23,958 | 24.1 |  | 6,043 | 14.7 |  | 3,872 | 14.6 |  | 2,857 | 23.0 |  | 7,877 | 23.0 |  | 209 | 32.4 |  | 12,209 | 31.7 |
| **Race and ethnicity** |  |  |  |  |  |  |  |  |  |  |  |  |  |  |  |  |  |  |  |  |  |  |  |
| Asian/Pacific Islander | 1,822 | 3.4 |  | 3,422 | 3.4 |  | 1,306 | 3.2 |  | 721 | 2.7 |  | 461 | 3.7 |  | 1,211 | 3.5 |  | 55 | 8.5 |  | 1,490 | 3.9 |
| Hispanic (any race) | 5,140 | 9.5 |  | 9,686 | 9.7 |  | 3,741 | 9.1 |  | 2,329 | 8.8 |  | 1,257 | 10.1 |  | 3,325 | 9.7 |  | 142 | 22.0 |  | 4,032 | 10.5 |
| Non-Hispanic Black | 10,437 | 19.3 |  | 19,596 | 19.7 |  | 7,803 | 19.0 |  | 5,055 | 19.0 |  | 2,485 | 20.0 |  | 6,914 | 20.2 |  | 149 | 23.1 |  | 7,627 | 19.8 |
| Non-Hispanic White | 36,167 | 66.8 |  | 65,469 | 65.9 |  | 27,812 | 67.7 |  | 18,149 | 68.4 |  | 8,066 | 65.0 |  | 22,446 | 65.4 |  | 289 | 44.8 |  | 24,874 | 64.6 |
| Other^a^ | 552 | 1.0 |  | 1,200 | 1.2 |  | 404 | 1.0 |  | 282 | 1.1 |  | 138 | 1.1 |  | 414 | 1.2 |  | 10 | 1.6 |  | 504 | 1.3 |
| **Transplanted organ^b^** | |  |  |  |  |  |  |  |  |  |  |  |  |  |  |  |  |  |  |  |  |  |  |
| Kidney only | 29,532 | 54.6 |  | 48,774 | 49.1 |  | 22,766 | 55.4 |  | 14,828 | 55.9 |  | 6,411 | 51.7 |  | 17,818 | 51.9 |  | 355 | 55.0 |  | 16,128 | 41.9 |
| Liver only | 11,856 | 21.9 |  | 23,134 | 23.3 |  | 8,753 | 21.3 |  | 5,585 | 21.0 |  | 2,976 | 24.0 |  | 7,992 | 23.3 |  | 127 | 19.7 |  | 9,557 | 24.8 |
| Lung only | 4,338 | 8.0 |  | 12,459 | 12.5 |  | 2,950 | 7.2 |  | 1,970 | 7.4 |  | 1,308 | 10.5 |  | 3,756 | 10.9 |  | 80 | 12.4 |  | 6,733 | 17.5 |
| Heart only | 5,336 | 9.9 |  | 9,530 | 9.6 |  | 4,253 | 10.4 |  | 2,725 | 10.3 |  | 1,039 | 8.4 |  | 2,914 | 8.5 |  | 44 | 6.8 |  | 3,891 | 10.1 |
| Other/multiple | 3,056 | 5.6 |  | 5,476 | 5.5 |  | 2,344 | 5.7 |  | 1,428 | 5.4 |  | 673 | 5.4 |  | 1,830 | 5.3 |  | 39 | 6.0 |  | 2,218 | 5.8 |
| **Years since transplant, attained** | | |  |  |  |  |  |  |  |  |  |  |  |  |  |  |  |  |  |  |  |  |  |
| <1 | 7,584 | 14.0 |  | 21,830 | 22.0 |  | 5,541 | 13.5 |  | 3,607 | 13.6 |  | 1,951 | 15.7 |  | 5,735 | 16.7 |  | 92 | 14.3 |  | 12,488 | 32.4 |
| 1–1.99 | 3,604 | 6.7 |  | 10,200 | 10.3 |  | 2,547 | 6.2 |  | 1,567 | 5.9 |  | 986 | 7.9 |  | 2,859 | 8.3 |  | 71 | 11.0 |  | 5,774 | 15.0 |
| 2–4.99 | 9,711 | 17.9 |  | 24,519 | 24.7 |  | 6,678 | 16.3 |  | 4,471 | 16.8 |  | 2,760 | 22.2 |  | 7,547 | 22.0 |  | 273 | 42.3 |  | 12,501 | 32.4 |
| 5–9.99 | 17,123 | 31.6 |  | 28,751 | 28.9 |  | 12,073 | 29.4 |  | 7,776 | 29.3 |  | 4,841 | 39.0 |  | 13,211 | 38.5 |  | 209 | 32.4 |  | 7,764 | 20.2 |
| ≥10 | 16,096 | 29.7 |  | 14,073 | 14.2 |  | 14,227 | 34.6 |  | 9,115 | 34.3 |  | 1,869 | 15.1 |  | 4,958 | 14.5 |  | 0 | 0.0 |  | 0 | 0.0 |

NDI, National Death Index; SRTR, Scientific Registry of Transplant Recipients; US, United States.

a. Includes races/ethnicities not already captured, and those with unknown or missing race/ethnicity information.
b. Single organ transplants of kidney, liver, lung, heart, and then other organs (pancreas, pancreas islets, intestine) and multiple transplants.

eTable 5. Frequency of underlying causes of death among adult organ recipients in the US during 1999–2019, by transplanted organ^a^

| **Cause of death** | **Overall** | |  | **Kidney only** | |  | **Liver only** | |  | **Lung only** | |  | **Heart only** | |  | **Pancreas ± kidney** | |  | **Other/**  **multiple** | |
| --- | --- | --- | --- | --- | --- | --- | --- | --- | --- | --- | --- | --- | --- | --- | --- | --- | --- | --- | --- | --- |
|  | **n** | **Col**  **%** |  | **n** | **Col**  **%** |  | **n** | **Col**  **%** |  | **n** | **Col**  **%** |  | **n** | **Col**  **%** |  | **n** | **Col**  **%** |  | **n** | **Col**  **%** |
| Heart disease | 25,945 | 16.9 |  | 16,273 | 20.8 |  | 2,448 | 7.0 |  | 787 | 4.6 |  | 5,215 | 35.1 |  | 766 | 16.5 |  | 455 | 14.6 |
| Graft failure | 22,875 | 14.9 |  | 5,381 | 6.9 |  | 7,780 | 22.3 |  | 5,642 | 33.2 |  | 3,280 | 22.1 |  | 24 | 0.5 |  | 768 | 9.4 |
| Cancer | 22,126 | 14.4 |  | 10,219 | 13.0 |  | 7,182 | 20.6 |  | 1,882 | 11.1 |  | 1,982 | 13.3 |  | 395 | 8.5 |  | 466 | 10.3 |
| Kidney diseases | 13,456 | 8.8 |  | 10,706 | 13.7 |  | 1,157 | 3.3 |  | 290 | 1.7 |  | 568 | 3.8 |  | 551 | 11.9 |  | 184 | 8.8 |
| Diabetes mellitus | 11,544 | 7.5 |  | 9,115 | 11.6 |  | 658 | 1.9 |  | 62 | 0.4 |  | 307 | 2.1 |  | 1,293 | 27.8 |  | 108 | 16.7 |
| Other infections | 10,365 | 6.8 |  | 4,541 | 5.8 |  | 3,904 | 11.2 |  | 670 | 4.0 |  | 662 | 4.5 |  | 271 | 5.8 |  | 316 | 7.0 |
| Liver diseases | 7,157 | 4.7 |  | 688 | 0.9 |  | 5,833 | 16.8 |  | 98 | 0.6 |  | 72 | 0.5 |  | 20 | 0.4 |  | 445 | 5.6 |
| Other respiratory diseases | 5,087 | 3.3 |  | 1,347 | 1.7 |  | 499 | 1.4 |  | 2,870 | 16.9 |  | 238 | 1.6 |  | 63 | 1.4 |  | 70 | 1.6 |
| Cerebrovascular diseases | 4,362 | 2.8 |  | 2,823 | 3.6 |  | 704 | 2.0 |  | 203 | 1.2 |  | 372 | 2.5 |  | 187 | 4.0 |  | 73 | 3.1 |
| Other circulatory diseases | 4,134 | 2.7 |  | 3,235 | 4.1 |  | 389 | 1.1 |  | 118 | 0.7 |  | 212 | 1.4 |  | 122 | 2.6 |  | 59 | 2.2 |
| Influenza and pneumonia | 3,215 | 2.1 |  | 1,891 | 2.4 |  | 400 | 1.2 |  | 524 | 3.1 |  | 237 | 1.6 |  | 99 | 2.1 |  | 64 | 2.0 |
| Accidents and adverse events | 3,143 | 2.1 |  | 1,777 | 2.3 |  | 792 | 2.3 |  | 116 | 0.7 |  | 235 | 1.6 |  | 134 | 2.9 |  | 89 | 2.7 |
| Chronic lower respiratory diseases | 3,030 | 2.0 |  | 994 | 1.3 |  | 349 | 1.0 |  | 1,531 | 9.0 |  | 122 | 0.8 |  | 15 | 0.3 |  | 19 | 0.4 |
| Dementia and Alzheimer’s | 818 | 0.5 |  | 615 | 0.8 |  | 94 | 0.3 |  | 18 | 0.1 |  | 69 | 0.5 |  | 21 | 0.4 |  | 2 | 0.3 |
| Cystic fibrosis | 771 | 0.5 |  | 10 | 0.0 |  | 12 | 0.0 |  | 709 | 4.2 |  | 0 | 0.0 |  | 1 | 0.0 |  | 39 | 0.5 |
| Suicide and self–inflicted injury | 702 | 0.5 |  | 361 | 0.5 |  | 197 | 0.6 |  | 29 | 0.2 |  | 40 | 0.3 |  | 58 | 1.3 |  | 16 | 0.9 |
| Other cause | 14,762 | 9.6 |  | 8,458 | 10.8 |  | 2,429 | 7.0 |  | 1,430 | 8.4 |  | 1,253 | 8.4 |  | 623 | 13.4 |  | 567 | 14.2 |
| Total^b^ | 153,491 | 100.0 |  | 78,434 | 100.0 |  | 34,827 | 100.0 |  | 16,979 | 100.0 |  | 14,866 | 100.0 |  | 4,644 | 100.0 |  | 3,741 | 100.0 |

Col, column; n, number; US, United States.

a. Single organ transplants of kidney, liver, lung, heart, and then other organs (pancreas, pancreas islets, intestine) and multiple transplants.
b. The 99,373 National Death Index linked deaths were weighted to represent 153,491 deaths among those receiving their first organ and dying during 1999–2019.

eTable 6. Frequency of underlying causes of death among adult organ recipients in the US in 2019, by transplanted organ^a^

| **Cause of death** | **Overall** | |  | **Kidney only** | |  | **Liver only** | |  | **Lung only** | |  | **Heart only** | |  | **Pancreas ± kidney** | |  | **Other/**  **multiple** | |
| --- | --- | --- | --- | --- | --- | --- | --- | --- | --- | --- | --- | --- | --- | --- | --- | --- | --- | --- | --- | --- |
|  | **n** | **Col**  **%** |  | **n** | **Col**  **%** |  | **n** | **Col**  **%** |  | **n** | **Col**  **%** |  | **n** | **Col**  **%** |  | **n** | **Col**  **%** |  | **n** | **Col**  **%** |
| Heart disease | 1,882 | 17.5 |  | 1,206 | 22.2 |  | 203 | 9.5 |  | 77 | 5.4 |  | 285 | 26.3 |  | 62 | 17.9 |  | 49 | 14.3 |
| Graft failure | 1,380 | 12.8 |  | 302 | 5.6 |  | 267 | 12.5 |  | 531 | 37.3 |  | 227 | 21.0 |  | 2 | 0.6 |  | 51 | 14.9 |
| Cancer | 1,783 | 16.6 |  | 822 | 15.2 |  | 527 | 24.6 |  | 184 | 12.9 |  | 166 | 15.3 |  | 40 | 11.6 |  | 45 | 13.2 |
| Kidney diseases | 859 | 8.0 |  | 645 | 11.9 |  | 77 | 3.6 |  | 31 | 2.2 |  | 48 | 4.4 |  | 41 | 11.9 |  | 17 | 4.9 |
| Diabetes mellitus | 709 | 6.6 |  | 531 | 9.8 |  | 66 | 3.1 |  | 9 | 0.6 |  | 27 | 2.5 |  | 67 | 19.3 |  | 8 | 2.5 |
| Other infections | 573 | 5.3 |  | 285 | 5.3 |  | 143 | 6.7 |  | 57 | 4.0 |  | 45 | 4.1 |  | 24 | 6.8 |  | 20 | 5.9 |
| Liver diseases | 427 | 4.0 |  | 41 | 0.7 |  | 321 | 15.0 |  | 8 | 0.6 |  | 8 | 0.7 |  | 1 | 0.3 |  | 48 | 14.1 |
| Other respiratory diseases | 388 | 3.6 |  | 113 | 2.1 |  | 41 | 1.9 |  | 196 | 13.8 |  | 16 | 1.5 |  | 14 | 4.1 |  | 8 | 2.3 |
| Cerebrovascular diseases | 331 | 3.1 |  | 187 | 3.5 |  | 59 | 2.8 |  | 26 | 1.8 |  | 32 | 3.0 |  | 17 | 4.9 |  | 9 | 2.8 |
| Other circulatory diseases | 261 | 2.4 |  | 195 | 3.6 |  | 27 | 1.3 |  | 14 | 1.0 |  | 14 | 1.3 |  | 6 | 1.8 |  | 5 | 1.6 |
| Influenza and pneumonia | 249 | 2.3 |  | 131 | 2.4 |  | 36 | 1.7 |  | 32 | 2.3 |  | 19 | 1.8 |  | 20 | 5.8 |  | 10 | 2.9 |
| Accidents and adverse events | 252 | 2.3 |  | 122 | 2.2 |  | 78 | 3.7 |  | 11 | 0.8 |  | 28 | 2.6 |  | 5 | 1.4 |  | 8 | 2.4 |
| Chronic lower respiratory diseases | 179 | 1.7 |  | 66 | 1.2 |  | 36 | 1.7 |  | 57 | 4.0 |  | 18 | 1.6 |  | 0 | 0.0 |  | 2 | 0.7 |
| Dementia and Alzheimer’s | 101 | 0.9 |  | 63 | 1.2 |  | 20 | 0.9 |  | 6 | 0.4 |  | 8 | 0.8 |  | 3 | 0.7 |  | 1 | 0.3 |
| Cystic fibrosis | 45 | 0.4 |  | 0 | 0.0 |  | 1 | 0.0 |  | 42 | 3.0 |  | 0 | 0.0 |  | 0 | 0.0 |  | 2 | 0.6 |
| Suicide and self–inflicted injury | 34 | 0.3 |  | 21 | 0.4 |  | 5 | 0.3 |  | 5 | 0.3 |  | 1 | 0.1 |  | 2 | 0.7 |  | 0 | 0.0 |
| Other cause | 1,294 | 12.0 |  | 689 | 12.7 |  | 228 | 10.7 |  | 136 | 9.6 |  | 140 | 13.0 |  | 43 | 12.3 |  | 57 | 16.6 |
| Total | 10,747 | 100.0 |  | 5,419 | 100.0 |  | 2,136 | 100.0 |  | 1,421 | 100.0 |  | 1,082 | 100.0 |  | 348 | 100.0 |  | 342 | 100.0 |

Col, column; n, number; US, United States.

a. Single organ transplants of kidney, liver, lung, heart, and pancreas with or without a kidney, and then other organs and multiple transplants.

### eTable 7. Distribution (column %) of underlying causes of death among adult organ recipients in the US, by year of death

| **Cause of death** | **1999** | **2000** | **2001** | **2002** | **2003** | **2004** | **2005** | **2006** | **2007** | **2008** | **2009** | **2010** | **2011** | **2012** | **2013** | **2014** | **2015** | **2016** | **2017** | **2018** | **2019** |
| --- | --- | --- | --- | --- | --- | --- | --- | --- | --- | --- | --- | --- | --- | --- | --- | --- | --- | --- | --- | --- | --- |
| Heart disease | 19.2 | 15.6 | 14.8 | 17.0 | 15.7 | 13.6 | 15.2 | 17.0 | 16.9 | 16.7 | 16.2 | 16.7 | 17.1 | 17.1 | 16.4 | 17.8 | 17.8 | 18.0 | 16.7 | 17.5 | 17.5 |
| Graft failure | 36.2 | 27.9 | 26.7 | 22.6 | 23.1 | 24.1 | 21.9 | 17.7 | 16.3 | 16.2 | 15.1 | 14.3 | 14.7 | 14.0 | 13.7 | 11.5 | 11.2 | 11.2 | 12.0 | 11.4 | 12.8 |
| Cancer | 1.8 | 4.3 | 7.2 | 7.4 | 10.0 | 10.0 | 11.8 | 12.1 | 13.5 | 12.8 | 13.9 | 14.1 | 15.8 | 16.6 | 15.0 | 16.0 | 16.3 | 15.3 | 16.6 | 16.3 | 16.6 |
| Kidney diseases | 7.2 | 7.0 | 7.4 | 7.0 | 9.0 | 8.1 | 8.0 | 8.9 | 9.2 | 8.8 | 9.3 | 9.0 | 8.5 | 7.9 | 8.8 | 9.0 | 9.8 | 9.3 | 9.1 | 9.3 | 8.0 |
| Diabetes mellitus | 4.5 | 4.1 | 6.6 | 6.1 | 6.3 | 8.7 | 7.5 | 8.4 | 7.0 | 7.5 | 7.3 | 7.8 | 8.1 | 7.9 | 8.4 | 8.3 | 8.2 | 8.1 | 7.1 | 6.5 | 6.6 |
| Other infections | 8.1 | 9.2 | 8.8 | 9.6 | 7.4 | 8.1 | 7.6 | 8.2 | 7.0 | 7.8 | 7.5 | 6.8 | 6.9 | 6.8 | 6.6 | 6.7 | 6.0 | 6.0 | 5.6 | 5.9 | 5.3 |
| Liver diseases | 4.5 | 7.4 | 4.2 | 5.9 | 4.6 | 4.9 | 4.9 | 5.4 | 5.5 | 4.7 | 5.3 | 5.2 | 4.5 | 4.5 | 4.7 | 4.1 | 4.5 | 4.3 | 4.6 | 4.1 | 4.0 |
| Other respiratory diseases | 2.9 | 4.0 | 3.8 | 4.1 | 2.8 | 2.9 | 3.2 | 2.9 | 3.0 | 3.1 | 3.3 | 3.5 | 2.9 | 3.2 | 2.9 | 3.5 | 3.3 | 3.5 | 3.7 | 3.7 | 3.6 |
| Cerebrovas-cular diseases | 2.9 | 2.7 | 1.9 | 2.1 | 2.7 | 2.3 | 2.8 | 2.6 | 3.1 | 2.4 | 2.5 | 3.0 | 2.6 | 2.6 | 2.9 | 2.9 | 3.3 | 3.0 | 3.1 | 3.2 | 3.1 |
| Other circulatory diseases | 1.3 | 2.4 | 1.6 | 2.3 | 2.0 | 2.1 | 2.3 | 2.6 | 3.1 | 2.7 | 3.3 | 2.8 | 3.2 | 2.5 | 2.7 | 2.9 | 2.9 | 2.8 | 2.8 | 2.8 | 2.4 |
| Influenza and pneumonia | 0.7 | 1.0 | 1.2 | 0.7 | 1.8 | 1.9 | 1.3 | 1.5 | 1.8 | 2.0 | 2.6 | 2.1 | 1.9 | 1.8 | 2.4 | 2.3 | 2.0 | 2.8 | 2.4 | 2.5 | 2.3 |
| Accidents and adverse events | 0.5 | 1.1 | 1.8 | 1.2 | 1.2 | 1.9 | 1.3 | 1.9 | 1.9 | 2.0 | 1.8 | 2.3 | 2.0 | 2.1 | 2.2 | 2.0 | 2.1 | 2.3 | 2.4 | 2.5 | 2.3 |
| Chronic lower respiratory diseases | 1.6 | 2.4 | 2.4 | 2.6 | 2.3 | 2.2 | 1.8 | 2.1 | 1.6 | 2.2 | 2.3 | 2.0 | 2.0 | 2.4 | 2.0 | 1.7 | 2.0 | 1.9 | 1.9 | 1.7 | 1.7 |
| Dementia and Alzheimer’s | 0.0 | 0.0 | 0.0 | 0.0 | 0.1 | 0.1 | 0.1 | 0.2 | 0.1 | 0.5 | 0.3 | 0.4 | 0.5 | 0.5 | 0.8 | 0.7 | 0.8 | 0.8 | 0.6 | 0.9 | 0.9 |
| Cystic fibrosis | 0.7 | 0.7 | 0.8 | 0.9 | 1.3 | 1.0 | 0.5 | 0.6 | 0.6 | 0.8 | 0.7 | 0.5 | 0.3 | 0.3 | 0.4 | 0.4 | 0.4 | 0.4 | 0.3 | 0.4 | 0.4 |
| Suicide and self-inflicted injury | 0.9 | 0.3 | 0.5 | 0.5 | 0.6 | 0.5 | 0.6 | 0.2 | 0.6 | 0.5 | 0.4 | 0.4 | 0.6 | 0.4 | 0.4 | 0.5 | 0.4 | 0.5 | 0.4 | 0.6 | 0.3 |
| Other cause | 7.2 | 9.9 | 10.4 | 10.1 | 9.2 | 7.8 | 9.2 | 7.9 | 8.8 | 9.5 | 8.4 | 9.2 | 8.5 | 9.5 | 9.7 | 9.6 | 9.0 | 9.8 | 10.8 | 11.1 | 12.0 |

### eTable 8. Causes of death among adult organ recipients with graft failure^a^ in the US during 1999–2019, by transplanted organ

| **Cause of death** | **Overall** | |  | **Kidney** | |  | **Liver** | |  | **Lung** | |  | **Heart** | |  | **Pancreas ± kidney** | |  | **Other/**  **multiple** | |
| --- | --- | --- | --- | --- | --- | --- | --- | --- | --- | --- | --- | --- | --- | --- | --- | --- | --- | --- | --- | --- |
|  | **n** | **Col**  **%** |  | **n** | **Col**  **%** |  | **n** | **Col %** |  | **n** | **Col**  **%** |  | **n** | **Col %** |  | **n** | **Col**  **%** |  | **n** | **Col**  **%** |
| Heart disease | 3,833 | 16.8 |  | 865 | 16.1 |  | 199 | 2.6 |  | 205 | 3.6 |  | 2,466 | 75.2 |  | 3 | 10.7 |  | 95 | 12.4 |
| Cancer | 1,788 | 7.8 |  | 474 | 8.8 |  | 929 | 11.9 |  | 274 | 4.9 |  | 60 | 1.8 |  | 0 | 0.0 |  | 51 | 6.6 |
| Kidney diseases | 1,691 | 7.4 |  | 1,312 | 24.4 |  | 153 | 2.0 |  | 91 | 1.6 |  | 92 | 2.8 |  | 3 | 10.7 |  | 41 | 5.3 |
| Diabetes mellitus | 761 | 3.3 |  | 640 | 11.9 |  | 40 | 0.5 |  | 16 | 0.3 |  | 46 | 1.4 |  | 6 | 27.1 |  | 12 | 1.6 |
| Other infections | 3,387 | 14.8 |  | 526 | 9.8 |  | 2,415 | 31.0 |  | 209 | 3.7 |  | 90 | 2.8 |  | 0 | 0.0 |  | 146 | 19.0 |
| Liver diseases | 3,306 | 14.5 |  | 88 | 1.6 |  | 3,005 | 38.6 |  | 23 | 0.4 |  | 12 | 0.4 |  | 0 | 0.0 |  | 177 | 23.1 |
| Other respiratory diseases | 2,239 | 9.8 |  | 80 | 1.5 |  | 51 | 0.7 |  | 2,056 | 36.4 |  | 18 | 0.6 |  | 0 | 0.0 |  | 33 | 4.3 |
| Cerebrovascular diseases | 246 | 1.1 |  | 115 | 2.1 |  | 55 | 0.7 |  | 39 | 0.7 |  | 30 | 0.9 |  | 0 | 0.0 |  | 8 | 1.0 |
| Other circulatory diseases | 485 | 2.1 |  | 309 | 5.8 |  | 102 | 1.3 |  | 21 | 0.4 |  | 41 | 1.2 |  | 0 | 0.0 |  | 13 | 1.7 |
| Influenza and pneumonia | 480 | 2.1 |  | 135 | 2.5 |  | 51 | 0.7 |  | 270 | 4.8 |  | 12 | 0.4 |  | 0 | 0.0 |  | 13 | 1.6 |
| Accidents and adverse events | 243 | 1.1 |  | 87 | 1.6 |  | 89 | 1.1 |  | 33 | 0.6 |  | 22 | 0.7 |  | 1 | 4.3 |  | 10 | 1.3 |
| Chronic lower respiratory diseases | 1,275 | 5.6 |  | 43 | 0.8 |  | 14 | 0.2 |  | 1,202 | 21.3 |  | 7 | 0.2 |  | 0 | 0.0 |  | 9 | 1.2 |
| Dementia and Alzheimer’s | 28 | 0.1 |  | 14 | 0.3 |  | 10 | 0.1 |  | 4 | 0.1 |  | 0 | 0.0 |  | 0 | 0.0 |  | 0 | 0.0 |
| Cystic fibrosis | 557 | 2.4 |  | 3 | 0.0 |  | 2 | 0.0 |  | 540 | 9.6 |  | 0 | 0.0 |  | 0 | 0.0 |  | 13 | 1.7 |
| Suicide and self-inflicted injury | 23 | 0.1 |  | 4 | 0.1 |  | 13 | 0.2 |  | 5 | 0.1 |  | 1 | 0.0 |  | 0 | 0.0 |  | 0 | 0.0 |
| Other cause | 2,532 | 11.1 |  | 684 | 12.7 |  | 652 | 8.4 |  | 655 | 11.6 |  | 382 | 11.6 |  | 11 | 47.2 |  | 148 | 19.2 |
| Total | 22,875 | 100.0 |  | 5,381 | 100.0 |  | 7,780 | 100.0 |  | 5,642 | 100.0 |  | 3,280 | 100.0 |  | 24 | 100.0 |  | 768 | 100.0 |

Col, column; US, United States.

a. We utilized SRTR variables that capture graft failure as an event (irrespective of death) and assumed that deaths occurring within 90 days of graft failure were deaths due to graft failure.

### eTable 9. Overall mortality and leading causes of death among organ recipients in the US, 1999–2019, according to demographic and transplant characteristics

| **Characteristics** | **Overall** | |  | **Heart disease** | |  | **Graft failure** | |  | **Cancer** | |
| --- | --- | --- | --- | --- | --- | --- | --- | --- | --- | --- | --- |
|  | **Deaths** | **MR^a^** |  | **Deaths** | **MR^a^** |  | **Deaths** | **MR^a^** |  | **Deaths** | **MR^a^** |
| **Total** | 153,491 | 4,259 |  | 25,945 | 720 |  | 22,875 | 635 |  | 22,126 | 614 |
| **Sex** |  |  |  |  |  |  |  |  |  |  |  |
| Female | 53,924 | 3,943 |  | 7,995 | 585 |  | 8,329 | 609 |  | 6,567 | 480 |
| Male | 99,567 | 4,451 |  | 17,950 | 803 |  | 14,546 | 650 |  | 15,559 | 696 |
| **Age at first transplant, years** | |  |  |  |  |  |  |  |  |  |  |
| 18–34 | 10,775 | 1,987 |  | 1,538 | 284 |  | 2,188 | 403 |  | 600 | 111 |
| 35–49 | 32,092 | 2,879 |  | 5,304 | 476 |  | 4,927 | 442 |  | 3,429 | 308 |
| 50–64 | 77,620 | 5,066 |  | 12,972 | 847 |  | 11,733 | 766 |  | 12,583 | 821 |
| 65–96 | 33,003 | 7,950 |  | 6,132 | 1477 |  | 4,027 | 970 |  | 5,514 | 1,328 |
| **Race and ethnicity** |  |  |  |  |  |  |  |  |  |  |  |
| Asian/Pacific Islander | 5,001 | 2,688 |  | 852 | 458 |  | 659 | 354 |  | 813 | 437 |
| Hispanic (any race) | 14,560 | 3,106 |  | 2,322 | 495 |  | 2,139 | 456 |  | 1,682 | 359 |
| Non-Hispanic Black | 30,047 | 4,271 |  | 6,146 | 874 |  | 4,095 | 582 |  | 2,846 | 405 |
| Non-Hispanic White | 102,089 | 4,625 |  | 16,318 | 739 |  | 15,746 | 713 |  | 16,602 | 752 |
| Other^b^ | 1795 | 4,632 |  | 307 | 792 |  | 236 | 609 |  | 182 | 470 |
| **Transplanted organ^c^** |  |  |  |  |  |  |  |  |  |  |  |
| Kidney only | 78,434 | 3,505 |  | 16,273 | 727 |  | 5,381 | 240 |  | 10,219 | 457 |
| Liver only | 34,827 | 4,830 |  | 2,448 | 340 |  | 7,780 | 1079 |  | 7,182 | 996 |
| Lung only | 16,979 | 11,507 |  | 787 | 533 |  | 5,642 | 3824 |  | 1,882 | 1,276 |
| Heart only | 14,866 | 5,392 |  | 5,215 | 1892 |  | 3,280 | 1190 |  | 1,982 | 719 |
| Other/multiple | 8,385 | 3,772 |  | 1,221 | 549 |  | 792 | 356 |  | 861 | 388 |
| **Calendar year of transplant** | |  |  |  |  |  |  |  |  |  |  |
| 1999–2004 | 67,602 | 4,720 |  | 12,496 | 873 |  | 8,743 | 610 |  | 8,733 | 610 |
| 2005–2009 | 46,717 | 4,267 |  | 7,746 | 707 |  | 6,788 | 620 |  | 7,319 | 668 |
| 2010–2014 | 27,744 | 3,668 |  | 4,148 | 548 |  | 4,876 | 645 |  | 4,622 | 611 |
| 2015–2019 | 11,428 | 3,564 |  | 1,555 | 485 |  | 2,468 | 770 |  | 1,452 | 453 |
| **Years since transplant, attained** | |  |  |  |  |  |  |  |  |  |  |
| <1 | 29,695 | 6,464 |  | 4,124 | 898 |  | 8,179 | 1,780 |  | 1,713 | 373 |
| 1–1.99 | 13,756 | 3,334 |  | 1,758 | 426 |  | 3,026 | 733 |  | 2,266 | 549 |
| 2–4.99 | 34,377 | 3,407 |  | 5,371 | 532 |  | 5,445 | 540 |  | 6,166 | 611 |
| 5–9.99 | 45,692 | 4,276 |  | 8,622 | 807 |  | 4,361 | 408 |  | 7,605 | 712 |
| ≥10 | 29,972 | 4,579 |  | 6,070 | 927 |  | 1,864 | 285 |  | 4,376 | 669 |

CI, confidence interval; MR, mortality rate; US, United States.

a. Deaths per 100,000 person-years.
b. Includes races and ethnicities not already captured, and individuals with unknown or missing race and ethnicity information.
c. Single organ transplants of kidney, liver, lung, heart, and then other organs (pancreas, pancreas islets, intestine) and multiple transplants.

### eTable 10. Associations of demographic and transplant characteristics with mortality from kidney diseases, diabetes, and infections, among adult organ recipients in the US, 1999–2019

|  | **Kidney diseases** | | |  | **Diabetes mellitus** | | |  | **Infections^c^** | | |
| --- | --- | --- | --- | --- | --- | --- | --- | --- | --- | --- | --- |
| **Characteristics** | **Deaths** | **MR^a^** | **MRR (95% CI)^b^** |  | **Deaths** | **MR^a^** | **MRR (95% CI)^b^** |  | **Deaths** | **MR^a^** | **MRR (95% CI)^b^** |
| **Total** | 13,456 | 373 | … |  | 11,544 | 320 | … |  | 10,365 | 288 | … |
| **Sex** |  |  |  |  |  |  |  |  |  |  |  |
| Female | 5,206 | 381 | 1 [Reference] |  | 4,128 | 302 | 1 [Reference] |  | 3,755 | 275 | 1 [Reference] |
| Male | 8,250 | 369 | 1.01 (0.97-1.06) |  | 7,416 | 332 | 1.16 (1.10-1.22) |  | 6,611 | 296 | 1.03 (0.98-1.09) |
| **Age at first transplant, years** | |  |  |  |  |  |  |  |  |  |  |
| 18–34 | 1,135 | 209 | 1 [Reference] |  | 816 | 151 | 1 [Reference] |  | 508 | 94 | 1 [Reference] |
| 35–49 | 3,006 | 270 | 1.41 (1.28-1.56) |  | 3,051 | 274 | 1.98 (1.77-2.21) |  | 2,474 | 222 | 2.16 (1.89-2.47) |
| 50–64 | 6,156 | 402 | 2.64 (2.41-2.89) |  | 5,640 | 368 | 3.65 (3.29-4.06) |  | 5,486 | 358 | 3.36 (2.95-3.82) |
| 65–96 | 3,159 | 761 | 5.38 (4.88-5.92) |  | 2,037 | 491 | 5.25 (4.69-5.88) |  | 1,898 | 457 | 5.03 (4.39-5.77) |
| **Race and ethnicity** |  |  |  |  |  |  |  |  |  |  |  |
| Asian/Pacific Islander | 467 | 251 | 0.65 (0.57-0.73) |  | 373 | 201 | 0.63 (0.55-0.72) |  | 415 | 223 | 0.93 (0.82-1.06) |
| Hispanic (any race) | 1,197 | 255 | 0.73 (0.67-0.79) |  | 1,478 | 315 | 1.06 (0.98-1.15) |  | 1,155 | 246 | 1.05 (0.97-1.14) |
| Non-Hispanic Black | 3,569 | 507 | 1.32 (1.25-1.40) |  | 2,608 | 371 | 1.12 (1.05-1.19) |  | 2,338 | 332 | 1.69 (1.58-1.80) |
| Non-Hispanic White | 8,076 | 366 | 1 [Reference] |  | 6,782 | 307 | 1 [Reference] |  | 6,350 | 288 | 1 [Reference] |
| Other^d^ | 148 | 381 | 1.01 (0.81-1.25) |  | 302 | 780 | 2.29 (1.96-2.68) |  | 107 | 277 | 1.17 (0.92-1.48) |
| **Transplanted organ^e^** |  |  |  |  |  |  |  |  |  |  |  |
| Kidney only | 10,707 | 478.3 | 1 [Reference] |  | 9,115 | 407 | 1 [Reference] |  | 4,541 | 203 | 1 [Reference] |
| Liver only | 1,157 | 160.4 | 0.32 (0.30-0.35) |  | 658 | 91 | 0.20 (0.18-0.23) |  | 3,904 | 541 | 2.66 (2.50-2.83) |
| Lung only | 90 | 61.1 | 0.42 (0.36-0.49) |  | 62 | 42 | 0.11 (0.08-0.15) |  | 670 | 454 | 2.24 (2.02-2.49) |
| Heart only | 568 | 206.2 | 0.38 (0.34-0.43) |  | 307 | 111 | 0.24 (0.20-0.28) |  | 662 | 240 | 1.12 (1.00-1.25) |
| Other/multiple | 735 | 330.6 | 0.89 (0.80-0.99) |  | 1,402 | 630 | 1.88 (1.74-2.04) |  | 587 | 264 | 1.67 (1.48-1.87) |
| **Calendar year of transplant** | |  |  |  |  |  |  |  |  |  |  |
| 1999–2004 | 6,774 | 473 | 1 [Reference] |  | 5,801 | 405 | 1 [Reference] |  | 4,657 | 325 | 1 [Reference] |
| 2005–2009 | 4,022 | 367 | 0.72 (0.69-0.76) |  | 3,494 | 319 | 0.72 (0.68-0.77) |  | 3,121 | 285 | 0.75 (0.71-0.80) |
| 2010–2014 | 1,967 | 260 | 0.53 (0.49-0.56) |  | 1,730 | 229 | 0.54 (0.50-0.58) |  | 1,796 | 237 | 0.54 (0.50-0.58) |
| 2015–2019 | 692 | 216 | 0.44 (0.40-0.48) |  | 519 | 162 | 0.43 (0.39-0.48) |  | 792 | 247 | 0.41 (0.37-0.45) |
| **Years since transplant, attained** | |  |  |  |  |  |  |  |  |  |  |
| <1 | 1,973 | 429 | 1 [Reference] |  | 1,329 | 289 | 1 [Reference] |  | 2,622 | 571 | 1 [Reference] |
| 1–1.99 | 873 | 212 | 0.48 (0.44-0.54) |  | 746 | 181 | 0.61 (0.54-0.68) |  | 1,067 | 259 | 0.45 (0.41-0.49) |
| 2–4.99 | 2,734 | 271 | 0.60 (0.55-0.65) |  | 2,574 | 255 | 0.82 (0.75-0.89) |  | 2,275 | 225 | 0.38 (0.35-0.41) |
| 5–9.99 | 4,474 | 419 | 0.89 (0.82-0.95) |  | 4,276 | 400 | 1.21 (1.11-1.32) |  | 2,684 | 251 | 0.40 (0.38-0.43) |
| ≥10 | 3,403 | 520 | 1.09 (1.00-1.18) |  | 2,620 | 400 | 1.16 (1.06-1.28) |  | 1,718 | 262 | 0.40 (0.37-0.44) |

CI, confidence interval; MR, mortality rate; MRR, mortality rate ratio; US, United States.

a. Deaths per 100,000 person–years.
b. MRRs are from multivariable models that include all the variables shown in the table.
c. Included all infections (viral, bacterial, fungal, and parasitic) except influenza and pneumonia which was classified as its own cause of death group.
d. Includes races and ethnicities not already captured, and individuals with unknown or missing race and ethnicity information.
e. Single organ transplants of kidney, liver, lung, heart, and then other organs (pancreas, pancreas islets, intestine) and multiple transplants.

### eTable 11. Associations of demographic and transplant characteristics with mortality from liver, other respiratory, and cerebrovascular diseases, among adult organ recipients in the US, 1999–2019

|  | **Liver diseases** | | |  | **Other respiratory diseases** | | |  | **Cerebrovascular diseases** | | |
| --- | --- | --- | --- | --- | --- | --- | --- | --- | --- | --- | --- |
| **Characteristics** | **Deaths** | **MR^a^** | **MRR (95% CI)^b^** |  | **Deaths** | **MR^a^** | **MRR (95% CI)^b^** |  | **Deaths** | **MR^a^** | **MRR (95% CI)^b^** |
| **Total** | 7,157 | 199 | … |  | 5,087 | 141 | … |  | 4,362 | 121 | … |
| **Sex** |  |  |  |  |  |  |  |  |  |  |  |
| Female | 2,566 | 188 | 1 [Reference] |  | 1,806 | 132 | 1 [Reference] |  | 1,667 | 122 | 1 [Reference] |
| Male | 4,591 | 205 | 0.96 (0.90-1.03) |  | 3,281 | 147 | 1.18 (1.09-1.27) |  | 2,695 | 120 | 0.97 (0.90-1.06) |
| **Age at first transplant, years** | |  |  |  |  |  |  |  |  |  |  |
| 18–34 | 434 | 80 | 1 [Reference] |  | 227 | 42 | 1 [Reference] |  | 231 | 43 | 1 [Reference] |
| 35–49 | 1,774 | 159 | 1.24 (1.07-1.42) |  | 683 | 61 | 1.63 (1.34-2.00) |  | 896 | 80 | 1.98 (1.61-2.44) |
| 50–64 | 3,863 | 252 | 1.47 (1.29-1.69) |  | 2,712 | 177 | 3.48 (2.90-4.17) |  | 2,194 | 143 | 4.04 (3.33-4.91) |
| 65–96 | 1,086 | 262 | 2.07 (1.78-2.40) |  | 1,466 | 353 | 6.69 (5.55-8.07) |  | 1,041 | 251 | 7.79 (6.36-9.55) |
| **Race and ethnicity** |  |  |  |  |  |  |  |  |  |  |  |
| Asian/Pacific Islander | 201 | 108 | 0.60 (0.50-0.73) |  | 123 | 66 | 0.77 (0.61-0.97) |  | 223 | 120 | 1.02 (0.85-1.22) |
| Hispanic (any race) | 920 | 196 | 1.02 (0.93-1.12) |  | 402 | 86 | 0.99 (0.87-1.13) |  | 468 | 100 | 0.93 (0.82-1.06) |
| Non-Hispanic Black | 780 | 111 | 1.14 (1.03-1.26) |  | 587 | 83 | 0.98 (0.87-1.09) |  | 896 | 127 | 1.15 (1.04-1.28) |
| Non-Hispanic White | 5,156 | 234 | 1 [Reference] |  | 3,923 | 178 | 1 [Reference] |  | 2,736 | 124 | 1 [Reference] |
| Other^c^ | 101 | 260 | 1.50 (1.17-1.92) |  | 53 | 138 | 1.38 (0.96-1.99) |  | 39 | 101 | 0.87 (0.59-1.28) |
| **Transplanted organ^d^** | |  |  |  |  |  |  |  |  |  |  |
| Kidney only | 688 | 31 | 1 [Reference] |  | 1,347 | 60 | 1 [Reference] |  | 2,823 | 126 | 1 [Reference] |
| Liver only | 5,833 | 809 | 25.4 (22.8-28.4) |  | 499 | 69 | 1.00 (0.87-1.15) |  | 704 | 98 | 0.70 (0.63-0.79) |
| Lung only | 98 | 67 | 1.90 (1.44-2.49) |  | 2,870 | 1,945 | 27.9 (25.5-30.6) |  | 203 | 138 | 1.06 (0.89-1.27) |
| Heart only | 72 | 26 | 0.81 (0.59-1.12) |  | 238 | 86 | 1.20 (1.00-1.44) |  | 372 | 135 | 0.94 (0.80-1.09) |
| Other/multiple | 465 | 209 | 7.38 (6.35-8.58) |  | 133 | 60 | 1.29 (1.02-1.63) |  | 260 | 117 | 1.23 (1.04-1.46) |
| **Calendar year of transplant** | |  |  |  |  |  |  |  |  |  |  |
| 1999–2004 | 2,721 | 190 | 1 [Reference] |  | 1,699 | 119 | 1 [Reference] |  | 2,038 | 142 | 1 [Reference] |
| 2005–2009 | 2,221 | 203 | 0.96 (0.89-1.04) |  | 1,566 | 143 | 0.90 (0.82-0.99) |  | 1,307 | 119 | 0.78 (0.71-0.86) |
| 2010–2014 | 1,388 | 183 | 0.76 (0.70-0.83) |  | 1,244 | 165 | 0.76 (0.68-0.84) |  | 746 | 99 | 0.65 (0.58-0.72) |
| 2015–2019 | 828 | 258 | 0.68 (0.61-0.75) |  | 578 | 180 | 0.54 (0.48-0.62) |  | 270 | 84 | 0.52 (0.44-0.60) |
| **Years since transplant, attained** | |  |  |  |  |  |  |  |  |  |  |
| <1 | 2,419 | 527 | 1 [Reference] |  | 1,419 | 309 | 1 [Reference] |  | 704 | 153 | 1 [Reference] |
| 1–1.99 | 695 | 168 | 0.32 (0.29-0.36) |  | 550 | 133 | 0.46 (0.40-0.51) |  | 303 | 73 | 0.48 (0.40-0.56) |
| 2–4.99 | 1,360 | 135 | 0.26 (0.24-0.28) |  | 1,191 | 118 | 0.44 (0.40-0.48) |  | 911 | 90 | 0.57 (0.51-0.65) |
| 5–9.99 | 1,572 | 147 | 0.27 (0.25-0.30) |  | 1,176 | 110 | 0.49 (0.44-0.55) |  | 1,370 | 128 | 0.81 (0.72-0.92) |
| ≥10 | 1,111 | 170 | 0.30 (0.27-0.34) |  | 751 | 115 | 0.71 (0.62-0.81) |  | 1,074 | 164 | 1.08 (0.94-1.25) |

CI, confidence interval; MR, mortality rate; MRR, mortality rate ratio; US, United States.

a. Deaths per 100,000 person–years.
b. MRRs are from multivariable models that include all the variables shown in the table.
c. Includes races and ethnicities not already captured, and individuals with unknown or missing race and ethnicity information.
d. Single organ transplants of kidney, liver, lung, heart, and then other organs (pancreas, pancreas islets, intestine) and multiple transplants.

### eTable 12. Associations of demographic and transplant characteristics with mortality from other circulatory diseases, influenza and pneumonia, and accidents and adverse events, among adult organ recipients in the US, 1999–2019

|  | **Other circulatory diseases** | | |  | **Influenza and pneumonia** | | |  | **Accidents and adverse events** | | |
| --- | --- | --- | --- | --- | --- | --- | --- | --- | --- | --- | --- |
| **Characteristics** | **Deaths** | **MR^a^** | **MRR (95% CI)^b^** |  | **Deaths** | **MR^a^** | **MRR (95% CI)^b^** |  | **Deaths** | **MR^a^** | **MRR (95% CI)^b^** |
| **Total** | 4,134 | 115 | … |  | 3,215 | 89 | … |  | 3,143 | 87 | … |
| **Sex** |  |  |  |  |  |  |  |  |  |  |  |
| Female | 1,541 | 113 | 1 [Reference] |  | 1,169 | 86 | 1 [Reference] |  | 971 | 71 | 1 [Reference] |
| Male | 2,593 | 116 | 1.08 (0.99-1.18) |  | 2,046 | 91 | 1.08 (0.98-1.19) |  | 2,172 | 97 | 1.33 (1.20-1.47) |
| **Age at first transplant, years** | |  |  |  |  |  |  |  |  |  |  |
| 18–34 | 384 | 71 | 1 [Reference] |  | 146 | 27 | 1 [Reference] |  | 336 | 62 | 1 [Reference] |
| 35–49 | 940 | 84 | 1.32 (1.12-1.57) |  | 522 | 47 | 1.89 (1.48-2.42) |  | 772 | 69 | 1.06 (0.88-1.26) |
| 50–64 | 1,900 | 124 | 2.42 (2.06-2.83) |  | 1,665 | 109 | 4.74 (3.77-5.96) |  | 1,402 | 91 | 1.44 (1.22-1.71) |
| 65–96 | 910 | 219 | 4.75 (4.00-5.64) |  | 882 | 213 | 10.09 (7.95-12.79) |  | 634 | 153 | 2.62 (2.18-3.15) |
| **Race and ethnicity** |  |  |  |  |  |  |  |  |  |  |  |
| Asian/Pacific Islander | 135 | 72 | 0.72 (0.57-0.91) |  | 163 | 88 | 1.14 (0.93-1.40) |  | 72 | 38 | 0.39 (0.29-0.52) |
| Hispanic (any race) | 483 | 103 | 1.13 (0.99-1.29) |  | 393 | 84 | 1.20 (1.03-1.39) |  | 288 | 61 | 0.64 (0.55-0.76) |
| Non-Hispanic Black | 1,328 | 189 | 1.92 (1.74-2.11) |  | 586 | 83 | 1.14 (1.01-1.29) |  | 461 | 66 | 0.73 (0.63-0.83) |
| Non-Hispanic White | 2,137 | 97 | 1 [Reference] |  | 2,037 | 92 | 1 [Reference] |  | 2,279 | 103 | 1 [Reference] |
| Other^c^ | 50 | 129 | 1.34 (0.92-1.95) |  | 36 | 94 | 1.21 (0.79-1.83) |  | 44 | 113 | 1.19 (0.81-1.75) |
| **Transplanted organ^d^** |  |  |  |  |  |  |  |  |  |  |  |
| Kidney only | 3,235 | 145 | 1 [Reference] |  | 1,891 | 84 | 1 [Reference] |  | 1,777 | 79 | 1 [Reference] |
| Liver only | 389 | 54 | 0.39 (0.34-0.45) |  | 400 | 55 | 0.59 (0.51-0.68) |  | 792 | 110 | 1.23 (1.10-1.38) |
| Lung only | 118 | 80 | 0.65 (0.51-0.82) |  | 524 | 355 | 4.15 (3.65-4.72) |  | 116 | 79 | 0.88 (0.69-1.12) |
| Heart only | 212 | 77 | 0.50 (0.42-0.61) |  | 237 | 86 | 0.88 (0.73-1.06) |  | 235 | 85 | 0.89 (0.74-1.08) |
| Other/multiple | 180 | 81 | 0.76 (0.62-0.93) |  | 163 | 74 | 1.24 (1.00-1.55) |  | 223 | 100 | 1.34 (1.11-1.63) |
| **Calendar year of transplant** | |  |  |  |  |  |  |  |  |  |  |
| 1999–2004 | 2,099 | 147 | 1 [Reference] |  | 1,348 | 94 | 1 [Reference] |  | 1,368 | 96 | 1 [Reference] |
| 2005–2009 | 1,200 | 110 | 0.69 (0.62-0.76) |  | 1,070 | 98 | 0.91 (0.81-1.02) |  | 938 | 86 | 0.89 (0.79-0.99) |
| 2010–2014 | 604 | 80 | 0.52 (0.46-0.58) |  | 574 | 76 | 0.67 (0.59-0.77) |  | 618 | 82 | 0.87 (0.77-0.99) |
| 2015–2019 | 231 | 72 | 0.50 (0.43-0.59) |  | 223 | 69 | 0.57 (0.47-0.68) |  | 219 | 68 | 0.75 (0.63-0.90) |
| **Years since transplant, attained** | |  |  |  |  |  |  |  |  |  |  |
| <1 | 545 | 119 | 1 [Reference] |  | 463 | 101 | 1 [Reference] |  | 391 | 85 | 1 [Reference] |
| 1–1.99 | 248 | 60 | 0.50 (0.42-0.60) |  | 303 | 73 | 0.74 (0.62-0.88) |  | 273 | 66 | 0.78 (0.64-0.94) |
| 2–4.99 | 833 | 83 | 0.68 (0.59-0.78) |  | 651 | 65 | 0.66 (0.57-0.77) |  | 764 | 76 | 0.89 (0.76-1.04) |
| 5–9.99 | 1,476 | 138 | 1.10 (0.96-1.26) |  | 1,043 | 98 | 1.05 (0.91-1.22) |  | 1,005 | 94 | 1.11 (0.95-1.31) |
| ≥10 | 1,031 | 158 | 1.23 (1.05-1.43) |  | 756 | 116 | 1.44 (1.22-1.71) |  | 710 | 109 | 1.33 (1.10-1.60) |

CI, confidence interval; MR, mortality rate; MRR, mortality rate ratio; US, United States.

a. Deaths per 100,000 person-years.
b. MRRs are from multivariable models that include all the variables shown in the table.
c. Includes races and ethnicities not already captured, and individuals with unknown or missing race and ethnicity information.
d. Single organ transplants of kidney, liver, lung, heart, and then other organs (pancreas, pancreas islets, intestine) and multiple transplants.

### eTable 13. Associations of demographic and transplant characteristics with mortality from chronic lower respiratory diseases, dementia and Alzheimer’s, and cystic fibrosis, among adult organ recipients in the US, 1999–2019

|  | **Chronic lower respiratory diseases** | | |  | **Dementia and Alzheimer’s** | | |  | **Cystic fibrosis** | | |
| --- | --- | --- | --- | --- | --- | --- | --- | --- | --- | --- | --- |
| **Characteristics** | **Deaths** | **MR^a^** | **MRR (95% CI)^b^** |  | **Deaths** | **MR^a^** | **MRR (95% CI)^b^** |  | **Deaths** | **MR^a^** | **MRR (95% CI)^b^** |
| **Total** | 3,030 | 84 | … |  | 818 | 0 | … |  | 771 | 21 | … |
| **Sex** |  |  |  |  |  |  |  |  |  |  |  |
| Female | 1,296 | 95 | 1 [Reference] |  | 353 | 26 | 1 [Reference] |  | 376 | 28 | 1 [Reference] |
| Male | 1,734 | 78 | 0.88 (0.80-0.97) |  | 465 | 21 | 0.80 (0.66-0.97) |  | 395 | 18 | 1.07 (0.89-1.30) |
| **Age at first transplant, years** | |  |  |  |  |  |  |  |  |  |  |
| 18–34 | 52 | 10 | 1 [Reference] |  | 4 | 1 | 1 [Reference] |  | 570 | 105 | 1 [Reference] |
| 35–49 | 335 | 30 | 3.55 (2.37-5.31) |  | 28 | 3 | 4.28 (0.85-21.47) |  | 168 | 15 | 0.20 (0.15-0.25) |
| 50–64 | 1,898 | 124 | 12.2 (8.3-17.7) |  | 337 | 22 | 49.1 (10.9-222) |  | 32 | 2 | 0.01 (0.01-0.02) |
| 65–96 | 745 | 179 | 21.7 (14.8-32.0) |  | 450 | 108 | 306 (68.3-1373) |  | 1 | 0 | 0.00 (0.00-0.01) |
| **Race and ethnicity** |  |  |  |  |  |  |  |  |  |  |  |
| Asian/Pacific Islander | 36 | 19 | 0.30 (0.20-0.47) |  | 28 | 15 | 0.56 (0.34-0.92) |  | 3 | 1 | 0.24 (0.03-1.72) |
| Hispanic (any race) | 153 | 33 | 0.55 (0.44-0.70) |  | 67 | 14 | 0.68 (0.49-0.94) |  | 16 | 3 | 0.29 (0.14-0.57) |
| Non-Hispanic Black | 273 | 39 | 0.63 (0.53-0.75) |  | 149 | 21 | 0.97 (0.76-1.24) |  | 14 | 2 | 0.21 (0.10-0.44) |
| Non-Hispanic White | 2,549 | 115 | 1 [Reference] |  | 571 | 26 | 1 [Reference] |  | 736 | 33 | 1 [Reference] |
| Other^c^ | 20 | 53 | 0.76 (0.42-1.37) |  | 4 | 9 | 0.40 (0.09-1.81) |  | 3 | 7 | 0.63 (0.09-4.56) |
| **Transplanted organ^d^** |  |  |  |  |  |  |  |  |  |  |  |
| Kidney only | 994 | 44 | 1 [Reference] |  | 615 | 27 | 1 [Reference] |  | 10 | 0 | 1 [Reference] |
| Liver only | 349 | 48 | 0.85 (0.71-1.00) |  | 94 | 13 | 0.41 (0.30-0.55) |  | 12 | 2 | 6.35 (1.94-20.8) |
| Lung only | 1,531 | 1,037 | 20.2 (18.0-22.6) |  | 18 | 12 | 0.48 (0.27-0.84) |  | 709 | 480 | 1103 (456-2668) |
| Heart only | 123 | 44 | 0.73 (0.56-0.95) |  | 69 | 25 | 0.70 (0.49-0.98) |  | 0 | 0 | 0.00 (0.00-0.00) |
| Other/multiple | 34 | 15 | 0.47 (0.29-0.76) |  | 23 | 10 | 0.87 (0.47-1.60) |  | 40 | 18 | 25.7 (9.6-68.6) |
| **Calendar year of transplant** | |  |  |  |  |  |  |  |  |  |  |
| 1999–2004 | 1,519 | 106 | 1 [Reference] |  | 367 | 26 | 1 [Reference] |  | 336 | 23 | 1 [Reference] |
| 2005–2009 | 895 | 82 | 0.59 (0.53-0.66) |  | 305 | 28 | 0.97 (0.79-1.19) |  | 248 | 23 | 0.89 (0.71-1.12) |
| 2010–2014 | 471 | 62 | 0.36 (0.32-0.42) |  | 130 | 17 | 0.82 (0.64-1.07) |  | 131 | 17 | 0.52 (0.40-0.67) |
| 2015–2019 | 146 | 46 | 0.22 (0.18-0.27) |  | 16 | 5 | 0.65 (0.36-1.16) |  | 56 | 17 | 0.40 (0.29-0.56) |
| **Years since transplant, attained** | |  |  |  |  |  |  |  |  |  |  |
| <1 | 454 | 99 | 1 [Reference] |  | 7 | 2 | 1 [Reference] |  | 198 | 43 | 1 [Reference] |
| 1–1.99 | 273 | 66 | 0.69 (0.57-0.84) |  | 23 | 5 | 3.65 (1.26-10.64) |  | 106 | 26 | 0.62 (0.46-0.84) |
| 2–4.99 | 686 | 68 | 0.73 (0.63-0.86) |  | 111 | 11 | 7.50 (2.78-20.24) |  | 226 | 22 | 0.58 (0.45-0.75) |
| 5–9.99 | 952 | 89 | 1.05 (0.90-1.23) |  | 357 | 33 | 25.6 (9.5-68.8) |  | 164 | 15 | 0.41 (0.31-0.55) |
| ≥10 | 665 | 102 | 1.40 (1.17-1.68) |  | 320 | 49 | 54.6 (20.0-149) |  | 76 | 12 | 0.31 (0.21-0.46) |

CI, confidence interval; MR, mortality rate; MRR, mortality rate ratio; US, United States.

a. Deaths per 100,000 person–years.
b. MRRs are from multivariable models that include all the variables shown in the table.
c. Includes races and ethnicities not already captured, and individuals with unknown or missing race and ethnicity information.
d. Single organ transplants of kidney, liver, lung, heart, and then other organs (pancreas, pancreas islets, intestine) and multiple transplants.

### eTable 14. Associations of demographic and transplant characteristics with mortality from suicide and self–inflicted injury and other causes among adult organ recipients in the US, 1999–2019

|  | **Suicide and self–inflicted injury** | | |  | **Other cause** | | |
| --- | --- | --- | --- | --- | --- | --- | --- |
| **Characteristics** | **Deaths** | **MR^a^** | **MRR (95% CI)^b^** |  | **Deaths** | **MR^a^** | **MRR (95% CI)^b^** |
| **Total** | 702 | 19 | … |  | 14,762 | 410 | … |
| **Sex** |  |  |  |  |  |  |  |
| Female | 127 | 9 | 1 [Reference] |  | 6,073 | 444 | 1 [Reference] |
| Male | 575 | 26 | 2.72 (2.09-3.54) |  | 8,689 | 388 | 0.87 (0.84-0.91) |
| **Age at first transplant, years** |  |  |  |  |  |  |  |
| 18–34 | 87 | 16 | 1 [Reference] |  | 1,520 | 280 | 1 [Reference] |
| 35–49 | 251 | 23 | 1.21 (0.86-1.71) |  | 3,533 | 317 | 1.17 (1.07-1.27) |
| 50–64 | 290 | 19 | 0.98 (0.69-1.38) |  | 6,759 | 441 | 1.75 (1.62-1.89) |
| 65–96 | 74 | 18 | 0.93 (0.62-1.40) |  | 2,950 | 711 | 3.02 (2.77-3.30) |
| **Race and ethnicity** |  |  |  |  |  |  |  |
| Asian/Pacific Islander | 19 | 10 | 0.45 (0.24-0.82) |  | 420 | 226 | 0.59 (0.52-0.67) |
| Hispanic (any race) | 53 | 11 | 0.46 (0.32-0.66) |  | 1,345 | 287 | 0.78 (0.72-0.84) |
| Non-Hispanic Black | 69 | 10 | 0.43 (0.31-0.60) |  | 3,303 | 470 | 1.26 (1.19-1.33) |
| Non-Hispanic White | 553 | 25 | 1 [Reference] |  | 9,539 | 432 | 1 [Reference] |
| Other^c^ | 9 | 22 | 0.95 (0.40-2.23) |  | 154 | 396 | 1.01 (0.82-1.23) |
| **Transplanted organ^d^** |  |  |  |  |  |  |  |
| Kidney only | 361 | 16 | 1 [Reference] |  | 8,458 | 378 | 1 [Reference] |
| Liver only | 197 | 27 | 1.40 (1.11-1.78) |  | 2,429 | 337 | 0.87 (0.82-0.93) |
| Lung only | 29 | 20 | 1.05 (0.65-1.68) |  | 1,430 | 969 | 2.45 (2.28-2.64) |
| Heart only | 40 | 15 | 0.71 (0.45-1.10) |  | 1,253 | 455 | 1.13 (1.04-1.22) |
| Other/multiple | 74 | 33 | 1.73 (1.21-2.46) |  | 1,191 | 536 | 1.66 (1.53-1.80) |
| **Calendar year of transplant** |  |  |  |  |  |  |  |
| 1999–2004 | 290 | 20 | 1 [Reference] |  | 6,613 | 462 | 1 [Reference] |
| 2005–2009 | 215 | 20 | 0.98 (0.77-1.26) |  | 4,263 | 389 | 0.81 (0.76-0.85) |
| 2010–2014 | 138 | 18 | 0.90 (0.68-1.18) |  | 2,558 | 338 | 0.69 (0.64-0.73) |
| 2015–2019 | 58 | 18 | 0.79 (0.55-1.12) |  | 1,327 | 414 | 0.71 (0.65-0.76) |
| **Years since transplant, attained** | |  |  |  |  |  |  |
| <1 | 121 | 26 | 1 [Reference] |  | 3,035 | 661 | 1 [Reference] |
| 1–1.99 | 82 | 20 | 0.75 (0.53-1.07) |  | 1,166 | 283 | 0.43 (0.40-0.47) |
| 2–4.99 | 196 | 19 | 0.71 (0.53-0.97) |  | 2,882 | 286 | 0.44 (0.41-0.47) |
| 5–9.99 | 179 | 17 | 0.59 (0.43-0.82) |  | 4,377 | 410 | 0.65 (0.61-0.69) |
| ≥10 | 124 | 19 | 0.64 (0.43-0.95) |  | 3,302 | 504 | 0.80 (0.74-0.86) |

CI, confidence interval; MR, mortality rate; MRR, mortality rate ratio; US, United States.

a. Deaths per 100,000 person-years.
b. MRRs are from multivariable models that include all the variables shown in the table.
c. Includes races and ethnicities not already captured, and individuals with unknown or missing race and ethnicity information.
d. Single organ transplants of kidney, liver, lung, heart, and then other organs (pancreas, pancreas islets, intestine) and multiple transplants.

### eTable 15. Associations of demographic and transplant characteristics with mortality from kidney diseases and diabetes mellitus among kidney recipients (N=85,413) in the US, 1999–2019

| **Characteristics** | **Kidney diseases** | | |  | **Diabetes mellitus** | | |  | **Graft failure** | | |
| --- | --- | --- | --- | --- | --- | --- | --- | --- | --- | --- | --- |
|  | **Deaths** | **MR^a^** | **MRR (95% CI)^b^** |  | **Deaths** | **MR^a^** | **MRR (95% CI)^b^** |  | **Deaths** | **MR^a^** | **MRR (95% CI)^b^** |
| **Total** | 11,405 | 469 | … |  | 10,434 | 429 | … |  | 5,878 | 242 | … |
| **Sex** |  |  |  |  |  |  |  |  |  |  |  |
| Female | 4,489 | 185 | 1 [Reference] |  | 3,791 | 156 | 1 [Reference] |  | 2,184 | 224 | 1 [Reference] |
| Male | 6,916 | 284 | 1.03 (0.98-1.08) |  | 6,643 | 273 | 1.16 (1.10-1.23) |  | 3,694 | 253 | 1.10 (1.03-1.18) |
| **Age at first transplant, years** |  |  |  |  |  |  |  |  |  |  |  |
| 18–34 | 1,070 | 44 | 1 [Reference] |  | 765 | 31 | 1 [Reference] |  | 283 | 64 | 1 [Reference] |
| 35–49 | 2,626 | 108 | 1.36 (1.23-1.51) |  | 2,843 | 117 | 2.06 (1.84-2.31) |  | 1,062 | 130 | 2.01 (1.67-2.42) |
| 50–64 | 4,967 | 204 | 2.59 (2.35-2.85) |  | 4,946 | 203 | 3.55 (3.18-3.96) |  | 2,873 | 322 | 5.07 (4.27-6.04) |
| 65–96 | 2,742 | 113 | 5.34 (4.82-5.91) |  | 1,880 | 77 | 4.95 (4.40-5.58) |  | 1,660 | 600 | 9.78 (8.17-11.7) |
| **Race and ethnicity** |  |  |  |  |  |  |  |  |  |  |  |
| Asian/Pacific Islander | 425 | 17 | 0.66 (0.58-0.75) |  | 355 | 15 | 0.60 (0.52-0.69) |  | 258 | 182 | 0.81 (0.69-0.95) |
| Hispanic (all races) | 1,053 | 43 | 0.73 (0.67-0.80) |  | 1,333 | 55 | 0.99 (0.91-1.07) |  | 651 | 188 | 0.92 (0.82-1.03) |
| Non-Hispanic Black | 3,323 | 137 | 1.32 (1.24-1.40) |  | 2,493 | 103 | 1.05 (0.98-1.12) |  | 1,625 | 276 | 1.28 (1.18-1.39) |
| No-Hispanic White | 6,471 | 266 | 1 [Reference] |  | 5,964 | 245 | 1 [Reference] |  | 3,242 | 245 | 1 [Reference] |
| Other^c^ | 133 | 5 | 1.00 (0.80-1.26) |  | 290 | 12 | 2.33 (1.98-2.73) |  | 103 | 349 | 1.48 (1.12-1.94) |
| **Calendar year of transplant** | |  |  |  |  |  |  |  |  |  |  |
| 1999–2004 | 5,778 | 238 | 1 [Reference] |  | 5,258 | 216 | 1 [Reference] |  | 2,627 | 265 | 1 [Reference] |
| 2005–2009 | 3,418 | 141 | 0.71 (0.67-0.75) |  | 3,143 | 129 | 0.72 (0.68-0.77) |  | 1,724 | 231 | 0.68 (0.62-0.74) |
| 2010–2014 | 1,653 | 68 | 0.51 (0.48-0.55) |  | 1,559 | 64 | 0.54 (0.51-0.58) |  | 1,096 | 221 | 0.52 (0.47-0.57) |
| 2015–2019 | 556 | 23 | 0.42 (0.38-0.47) |  | 475 | 20 | 0.45 (0.40-0.50) |  | 432 | 217 | 0.35 (0.31-0.40) |
| **Years since transplant, attained** | |  |  |  |  |  |  |  |  |  |  |
| <1 | 1,676 | 69 | 1 [Reference] |  | 1,217 | 50 | 1 [Reference] |  | 1,798 | 609 | 1 [Reference] |
| 1–1.99 | 724 | 30 | 0.47 (0.42-0.52) |  | 680 | 28 | 0.60 (0.54-0.68) |  | 529 | 196 | 0.32 (0.28-0.36) |
| 2–4.99 | 2,374 | 98 | 0.60 (0.55-0.65) |  | 2,337 | 96 | 0.81 (0.74-0.88) |  | 1,357 | 201 | 0.32 (0.29-0.35) |
| 5–9.99 | 3,845 | 158 | 0.87 (0.80-0.94) |  | 3,871 | 159 | 1.19 (1.09-1.30) |  | 1,494 | 203 | 0.31 (0.29-0.35) |
| ≥10 | 2,785 | 115 | 1.01 (0.93-1.11) |  | 2,329 | 96 | 1.12 (1.02-1.24) |  | 701 | 153 | 0.25 (0.21-0.28) |

CI, confidence interval; MR, mortality rate; MRR, mortality rate ratio; US, United States.

a. Deaths per 100,000 person-years.
b. MRRs are from multivariable models that include all the variables shown in the table.
c. Includes races and ethnicities not already captured, and individuals with unknown or missing race and ethnicity information.

### eTable 16. Associations of demographic and transplant characteristics with mortality from liver diseases and graft failure among adult liver recipients (N=37,373) in the US, 1999–2019

| **Characteristics** | **Liver diseases** | | |  | **Graft failure** | | |
| --- | --- | --- | --- | --- | --- | --- | --- |
|  | **Deaths** | **MR^a^** | **MRR (95% CI)^b^** |  | **Deaths** | **MR^a^** | **MRR (95% CI)^b^** |
| **Total** | 6,280 | 822 | … |  | 8,286 | 1,086 | … |
| **Sex** |  |  |  |  |  |  |  |
| Female | 2,300 | 301 | 1 [Reference] |  | 2,777 | 1,064 | 1 [Reference] |
| Male | 3,971 | 520 | 0.91 (0.85-0.97) |  | 5,510 | 1,097 | 1.04 (0.98-1.11) |
| **Age at first transplant, years** | |  |  |  |  |  |  |
| 18–34 | 368 | 48 | 1 [Reference] |  | 523 | 1,092 | 1 [Reference] |
| 35–49 | 1,557 | 204 | 1.04 (0.89-1.21) |  | 2,162 | 1,080 | 1.00 (0.88-1.13) |
| 50–64 | 3,419 | 448 | 1.05 (0.91-1.21) |  | 4,655 | 1,075 | 1.01 (0.90-1.14) |
| 65–96 | 927 | 121 | 1.46 (1.25-1.71) |  | 946 | 1,151 | 1.09 (0.94-1.25) |
| **Race and ethnicity** |  |  |  |  |  |  |  |
| Asian/Pacific Islander | 146 | 19 | 0.49 (0.40-0.61) |  | 263 | 741 | 0.75 (0.64-0.89) |
| Hispanic (any races) | 789 | 103 | 0.95 (0.86-1.05) |  | 1,004 | 1,000 | 1.00 (0.92-1.09) |
| Non-Hispanic Black | 616 | 81 | 1.21 (1.09-1.35) |  | 1,148 | 1,873 | 1.81 (1.66-1.97) |
| Non-Hispanic White | 4,640 | 608 | 1 [Reference] |  | 5,769 | 1,034 | 1 [Reference] |
| Other^c^ | 80 | 10 | 1.18 (0.90-1.54) |  | 103 | 1,256 | 1.25 (0.98-1.60) |
| **Calendar year of transplant** | |  |  |  |  |  |  |
| 1999–2004 | 2,336 | 306 | 1 [Reference] |  | 3,518 | 1,235 | 1 [Reference] |
| 2005–2009 | 1,913 | 251 | 0.94 (0.86-1.02) |  | 2,531 | 1,079 | 0.71 (0.66-0.76) |
| 2010–2014 | 1,243 | 163 | 0.75 (0.68-0.82) |  | 1,496 | 896 | 0.43 (0.40-0.47) |
| 2015–2019 | 778 | 102 | 0.65 (0.59-0.72) |  | 741 | 962 | 0.26 (0.24-0.29) |
| **Years since transplant, attained** | |  |  |  |  |  |  |
| <1 | 2,302 | 302 | 1 [Reference] |  | 4,088 | 3,966 | 1 [Reference] |
| 1–1.99 | 643 | 84 | 0.32 (0.28-0.35) |  | 1,175 | 1,304 | 0.32 (0.29-0.35) |
| 2–4.99 | 1,164 | 152 | 0.23 (0.21-0.26) |  | 1,629 | 762 | 0.17 (0.16-0.19) |
| 5–9.99 | 1,273 | 167 | 0.24 (0.21-0.26) |  | 1,033 | 467 | 0.09 (0.08-0.10) |
| ≥10 | 889 | 117 | 0.25 (0.23-0.29) |  | 362 | 267 | 0.04 (0.04-0.05) |

CI, confidence interval; MR, mortality rate; MRR, mortality rate ratio; US, United States.

a. Deaths per 100,000 person–years.
b. MRRs are from multivariable models that include all the variables shown in the table.
c. Includes races and ethnicities not already captured, and individuals with unknown or missing race and ethnicity information.

### eTable 17. Associations of demographic and transplant characteristics with mortality from chronic lower and other respiratory system disease and influenza and pneumonia, among lung recipients (N=17,376) in the US, 1999–2019

| **Characteristics** | **Chronic lower respiratory**  **system diseases** | | |  | **Other respiratory diseases** | | |  | **Influenza and pneumonia** | | |
| --- | --- | --- | --- | --- | --- | --- | --- | --- | --- | --- | --- |
|  | **Deaths** | **MR^a^** | **MRR (95% CI)^b^** |  | **Deaths** | **MR^a^** | **MRR (95% CI)^b^** |  | **Deaths** | **MR^a^** | **MRR (95% CI)^b^** |
| **Total** | 1,534 | 989 | … |  | 2,892 | 1,913 | … |  | 528 | 349 | … |
| **Sex** |  |  |  |  |  |  |  |  |  |  |  |
| Female | 774 | 776 | 1 [Reference] |  | 1,062 | 1,579 | 1 [Reference] |  | 237 | 352 | 1 [Reference] |
| Male | 760 | 1,161 | 0.75 (0.66-0.86) |  | 1,830 | 2,180 | 1.26 (1.14-1.38) |  | 291 | 347 | 0.94 (0.75-1.17) |
| **Age at first transplant, years** | |  |  |  |  |  |  |  |  |  |  |
| 18–34 | 33 | 98 | 1 [Reference] |  | 127 | 657 | 1 [Reference] |  | 44 | 227 | 1 [Reference] |
| 35–49 | 144 | 675 | 2.91 (1.76-4.82) |  | 365 | 1,288 | 1.95 (1.50-2.54) |  | 69 | 244 | 1.10 (0.66-1.82) |
| 50–64 | 1,050 | 1,082 | 8.43 (5.37-13.2) |  | 1,591 | 2,029 | 3.03 (2.40-3.83) |  | 304 | 388 | 1.79 (1.18-2.70) |
| 65–96 | 308 | 1,737 | 11.7 (7.33-18.7) |  | 809 | 3,214 | 4.87 (3.83-6.19) |  | 111 | 440 | 2.33 (1.49-3.63) |
| **Race and ethnicity** |  |  |  |  |  |  |  |  |  |  |  |
| Asian/Pacific Islander | 11 | 1,162 | 0.54 (0.26-1.14) |  | 56 | 2,601 | 1.37 (1.01-1.87) |  | 5 | 222 | 0.67 (0.25-1.82) |
| Hispanic (any race) | 42 | 1,393 | 0.65 (0.43-0.99) |  | 188 | 2,373 | 1.39 (1.17-1.66) |  | 37 | 469 | 1.49 (0.97-2.31) |
| Non-Hispanic Black | 92 | 1,463 | 0.78 (0.60-1.01) |  | 223 | 1,800 | 1.09 (0.92-1.28) |  | 31 | 251 | 0.77 (0.49-1.22) |
| Non-Hispanic White | 1,380 | 913 | 1 [Reference] |  | 2,409 | 1,884 | 1 [Reference] |  | 450 | 352 | 1 [Reference] |
| Other^c^ | 9 | 1,308 | 1.04 (0.44-2.48) |  | 16 | 1,900 | 1.11 (0.60-2.05) |  | 5 | 595 | 1.75 (0.64-4.75) |
| **Calendar year of transplant** | |  |  |  |  |  |  |  |  |  |  |
| 1999–2004 | 713 | 1,945 | 1 [Reference] |  | 777 | 1,878 | 1 [Reference] |  | 166 | 400 | 1 [Reference] |
| 2005–2009 | 455 | 998 | 0.51 (0.43-0.59) |  | 867 | 1,968 | 0.87 (0.76-1.00) |  | 177 | 402 | 0.91 (0.68-1.23) |
| 2010–2014 | 258 | 499 | 0.26 (0.21-0.31) |  | 822 | 1,928 | 0.70 (0.61-0.80) |  | 131 | 308 | 0.62 (0.46-0.85) |
| 2015–2019 | 108 | 167 | 0.16 (0.13-0.21) |  | 426 | 1,841 | 0.47 (0.40-0.55) |  | 54 | 233 | 0.36 (0.25-0.53) |
| **Years since transplant, attained** | | |  |  |  |  |  |  |  |  |  |
| <1 | 394 | 1,367 | 1 [Reference] |  | 1,096 | 3,801 | 1 [Reference] |  | 144 | 500 | 1 [Reference] |
| 1–1.99 | 213 | 903 | 0.64 (0.51-0.80) |  | 407 | 1,723 | 0.45 (0.39-0.51) |  | 101 | 427 | 0.83 (0.61-1.13) |
| 2–4.99 | 430 | 885 | 0.57 (0.47-0.69) |  | 749 | 1,541 | 0.38 (0.34-0.43) |  | 131 | 270 | 0.49 (0.36-0.66) |
| 5–9.99 | 390 | 1,064 | 0.59 (0.48-0.72) |  | 474 | 1,294 | 0.31 (0.27-0.36) |  | 107 | 291 | 0.48 (0.35-0.66) |
| ≥10 | 107 | 789 | 0.37 (0.27-0.51) |  | 166 | 1,228 | 0.31 (0.25-0.40) |  | 45 | 335 | 0.54 (0.33-0.88) |

CI, confidence interval; MR, mortality rate; MRR, mortality rate ratio; US, United States.

a. Deaths per 100,000 person-years.
b. MRRs are from multivariable models that include all the variables shown in the table.
c. Includes races and ethnicities not already captured, and individuals with unknown or missing race and ethnicity information.

### eTable 18. Associations of demographic and transplant characteristics with mortality from cystic fibrosis and graft failure among adult lung recipients (N=17,376) in the US, 1999–2019

| **Characteristics** | **Cystic fibrosis** | | |  |  | **Graft failure** | | |
| --- | --- | --- | --- | --- | --- | --- | --- | --- |
|  | **Deaths** | **MR^a^** | **MRR (95% CI)^b^** |  |  | **Deaths** | **MR^a^** | **MRR (95% CI)^b^** |
| **Total** | 745 | 493 | … |  |  | 5,784 | 3,825 | … |
| **Sex** |  |  |  |  |  |  |  |  |
| Female | 365 | 543 | 1 [Reference] |  |  | 2,555 | 3,799 | 1 [Reference] |
| Male | 380 | 453 | 1.11 (0.91-1.34) |  |  | 3,229 | 3,847 | 1.00 (0.93-1.06) |
| **Age at first transplant, years** |  |  |  |  |  |  |  |  |
| 18–34 | 550 | 2,851 | 1 [Reference] |  |  | 771 | 4,000 | Reference |
| 35–49 | 162 | 572 | 0.21 (0.17-0.27) |  |  | 917 | 3,235 | 0.80 (0.71-0.90) |
| 50–64 | 32 | 41 | 0.01 (0.01-0.02) |  |  | 2,953 | 3,766 | 0.92 (0.84-1.02) |
| 65–96 | 1 | 4 | 0.00 (0.00-0.01) |  |  | 1,144 | 4,541 | 1.10 (0.98-1.23) |
| **Race and ethnicity** |  |  |  |  |  |  |  |  |
| Asian/Pacific Islander | 3 | 119 | 0.26 (0.04-1.82) |  |  | 84 | 3,920 | 1.02 (0.80-1.30) |
| Hispanic (any race) | 16 | 199 | 0.31 (0.16-0.60) |  |  | 286 | 3,603 | 0.95 (0.83-1.09) |
| Non-Hispanic Black | 13 | 105 | 0.24 (0.11-0.52) |  |  | 535 | 4,314 | 1.18 (1.06-1.32) |
| Non-Hispanic White | 711 | 556 | 1 [Reference] |  |  | 4,852 | 3,794 | 1 [Reference] |
| Other^c^ | 3 | 297 | 0.57 (0.08-4.07) |  |  | 28 | 3,225 | 0.85 (0.56-1.29) |
| **Calendar year of transplant** | |  |  |  |  |  |  |  |
| 1999–2004 | 321 | 776 | 1 [Reference] |  |  | 1,485 | 3,589 | 1 [Reference] |
| 2005–2009 | 242 | 550 | 0.92 (0.73-1.15) |  |  | 1,642 | 3,727 | 0.99 (0.90-1.10) |
| 2010–2014 | 127 | 298 | 0.52 (0.40-0.67) |  |  | 1,732 | 4,064 | 1.02 (0.92-1.13) |
| 2015–2019 | 55 | 237 | 0.40 (0.29-0.57) |  |  | 925 | 3,998 | 0.94 (0.84-1.06) |
| **Years since transplant, attained** |  |  |  |  |  |  |  |  |
| <1 | 195 | 675 | 1 [Reference] |  |  | 1,261 | 4,373 | 1 [Reference] |
| 1–1.99 | 103 | 434 | 0.61 (0.45-0.83) |  |  | 957 | 4,049 | 0.93 (0.84-1.02) |
| 2–4.99 | 222 | 458 | 0.59 (0.46-0.76) |  |  | 1,835 | 3,777 | 0.86 (0.79-0.94) |
| 5–9.99 | 153 | 418 | 0.41 (0.30-0.55) |  |  | 1,320 | 3,601 | 0.83 (0.75-0.92) |
| ≥10 | 73 | 537 | 0.31 (0.21-0.47) |  |  | 411 | 3,046 | 0.72 (0.62-0.85) |

CI, confidence interval; MR, mortality rate; MRR, mortality rate ratio; US, United States.

a. Deaths per 100,000 person-years.
b. MRRs are from multivariable models that include all the variables shown in the table.
c. Includes races and ethnicities not already captured, and individuals with unknown or missing race and ethnicity information.

### eTable 19. Associations of demographic and transplant characteristics with mortality from heart disease and graft failure among adult heart recipients (N=15,625) in the US, 1999–2019

| **Characteristics** | **Heart disease** | | |  | **Graft failure** | | |
| --- | --- | --- | --- | --- | --- | --- | --- |
|  | **Deaths** | **MR^a^** | **MRR (95% CI)^b^** |  | **Deaths** | **MR^a^** | **MRR (95% CI)^b^** |
| **Total** | 5,428 | 1,894 | … |  | 3,460 | 1,207 | … |
| **Sex** |  |  |  |  |  |  |  |
| Female | 1,277 | 1,822 | 1 [Reference] |  | 976 | 1,392 | 1 [Reference] |
| Male | 4,151 | 1,917 | 1.05 (0.96-1.14) |  | 2,484 | 1,147 | 0.94 (0.86-1.04) |
| **Age at first transplant, years** |  |  |  |  |  |  |  |
| 18–34 | 545 | 1,789 | 1 [Reference] |  | 659 | 2,163 | 1 [Reference] |
| 35–49 | 1,109 | 1,679 | 0.94 (0.82-1.08) |  | 906 | 1,373 | 0.65 (0.57-0.74) |
| 50–64 | 2,870 | 1,881 | 1.08 (0.95-1.22) |  | 1,544 | 1,012 | 0.50 (0.45-0.57) |
| 65–96 | 905 | 2,405 | 1.45 (1.25-1.68) |  | 351 | 932 | 0.46 (0.39-0.54) |
| **Race and ethnicity** |  |  |  |  |  |  |  |
| Asian/Pacific Islander | 124 | 1,509 | 0.90 (0.72-1.13) |  | 84 | 1,023 | 0.90 (0.70-1.15) |
| Hispanic (any race) | 382 | 1,836 | 1.07 (0.93-1.24) |  | 269 | 1,293 | 1.12 (0.96-1.32) |
| Non-Hispanic Black | 1,075 | 2,294 | 1.36 (1.24-1.49) |  | 907 | 1,935 | 1.62 (1.46-1.79) |
| Non-Hispanic White | 3,813 | 1,828 | 1 [Reference] |  | 2,177 | 1,043 | 1 [Reference] |
| Other^c^ | 35 | 1,587 | 0.95 (0.62-1.46) |  | 23 | 1,068 | 0.93 (0.56-1.53) |
| **Calendar year of transplant** | |  |  |  |  |  |  |
| 1999–2004 | 2,647 | 2,284 | 1 [Reference] |  | 1,236 | 1,066 | 1 [Reference] |
| 2005–2009 | 1,344 | 1,669 | 0.69 (0.63-0.75) |  | 1,066 | 1,324 | 1.13 (1.01-1.27) |
| 2010–2014 | 889 | 1,450 | 0.53 (0.48-0.58) |  | 689 | 1,125 | 0.82 (0.73-0.93) |
| 2015–2019 | 549 | 1,895 | 0.45 (0.40-0.51) |  | 469 | 1,618 | 0.74 (0.65-0.85) |
| **Years since transplant, attained** | |  |  |  |  |  |  |
| <1 | 1,849 | 4,809 | 1 [Reference] |  | 1,301 | 3,382 | 1 [Reference] |
| 1–1.99 | 508 | 1,495 | 0.31 (0.27-0.35) |  | 443 | 1,304 | 0.38 (0.33-0.44) |
| 2–4.99 | 1,001 | 1,234 | 0.24 (0.22-0.27) |  | 728 | 897 | 0.26 (0.23-0.29) |
| 5–9.99 | 1,201 | 1,441 | 0.26 (0.23-0.29) |  | 589 | 707 | 0.19 (0.17-0.22) |
| ≥10 | 869 | 1,747 | 0.27 (0.24-0.31) |  | 399 | 802 | 0.21 (0.18-0.25) |

CI, confidence interval; MR, mortality rate; MRR, mortality rate ratio; US, United States.

a. Deaths per 100,000 person-years.
b. MRRs are from multivariable models that include all the variables shown in the table.
c. Includes races and ethnicities not already captured, and individuals with unknown or missing race and ethnicity information.

### eTable 20. Deaths and mortality rates among adult organ recipients in the US, by calendar period

| **Cause of death^a^** | **1999–2019** | |  | **1999–2004** | |  | **2005–2009** | |  | **2010–2014** | |  | **2015–2019** | |
| --- | --- | --- | --- | --- | --- | --- | --- | --- | --- | --- | --- | --- | --- | --- |
|  | **Deaths^b^** | **MR^c^** |  | **Deaths** | **MR^c^** |  | **Deaths** | **MR^c^** |  | **Deaths** | **MR^c^** |  | **Deaths** | **MR^c^** |
| Overall | 153,491 | 4,259 |  | 16,276 | 5,268 |  | 33,129 | 4,557 |  | 48,481 | 4,410 |  | 55,604 | 3,786 |
| Heart disease | 29,778 | 826 |  | 3,126 | 1,012 |  | 6,402 | 881 |  | 9,361 | 851 |  | 10,890 | 741 |
| Cancer | 23,914 | 664 |  | 1,534 | 496 |  | 4,728 | 650 |  | 8,058 | 733 |  | 9,595 | 653 |
| Kidney diseases | 15,146 | 420 |  | 1,618 | 524 |  | 3,387 | 466 |  | 4,647 | 423 |  | 5,495 | 374 |
| Diabetes mellitus | 12,305 | 341 |  | 1,210 | 392 |  | 2,632 | 362 |  | 4,195 | 382 |  | 4,268 | 291 |
| Other infections^d^ | 13,752 | 382 |  | 2,267 | 734 |  | 3,589 | 494 |  | 4,219 | 384 |  | 3,677 | 250 |
| Liver diseases | 10,463 | 290 |  | 1,689 | 547 |  | 2,563 | 353 |  | 3,133 | 285 |  | 3,078 | 210 |
| Other respiratory diseases | 7,326 | 203 |  | 688 | 223 |  | 1,456 | 200 |  | 2,268 | 206 |  | 2,914 | 198 |
| Cerebrovascular diseases | 4,608 | 128 |  | 438 | 142 |  | 948 | 130 |  | 1,418 | 129 |  | 1,804 | 123 |
| Other circulatory diseases | 4,619 | 128 |  | 425 | 138 |  | 1,053 | 145 |  | 1,505 | 137 |  | 1,635 | 111 |
| Influenza and pneumonia | 3,695 | 103 |  | 280 | 91 |  | 728 | 100 |  | 1,153 | 105 |  | 1,534 | 104 |
| Accidents and adverse events | 3,386 | 94 |  | 296 | 96 |  | 655 | 90 |  | 1,068 | 97 |  | 1,368 | 93 |
| Chronic lower respiratory diseases | 4,306 | 119 |  | 512 | 166 |  | 940 | 129 |  | 1,371 | 125 |  | 1,483 | 101 |
| Dementia and Alzheimer’s | 846 | 23 |  | 5 | 2 |  | 85 | 12 |  | 298 | 27 |  | 458 | 31 |
| Cystic fibrosis | 1,329 | 37 |  | 209 | 68 |  | 351 | 48 |  | 370 | 34 |  | 398 | 27 |
| Suicide and self-inflicted injury | 725 | 20 |  | 89 | 29 |  | 153 | 21 |  | 232 | 21 |  | 250 | 17 |
| Other cause of death | 17,294 | 480 |  | 1,890 | 612 |  | 3,461 | 476 |  | 5,185 | 472 |  | 6,757 | 460 |

MR, mortality rate; US, United States.

a. Since there is no International Classification of Diseases 10th Revision (ICD-10) code for graft failure, deaths classified as due to graft failure in the primary analyses were recategorized into the 16 cause of death groups accordingly to their NDI-assigned ICD-10 codes.
b. The 99,373 National Death Index linked deaths were weighted to represent 153,491 deaths among those receiving their first organ and dying during 1999–2019.
c. Deaths per 100,000 person-years.
d. This category includes all infections (viral, bacterial, fungal, and parasitic) except influenza and pneumonia which are classified as a separate cause of death group.

### eTable 21. Deaths and mortality rates among adult organ recipients in the US deaths during 2015–2019, by transplanted organ^a^

| **Cause of death^b^** | **Kidney** | |  | **Liver** | |  | **Lung** | |  | **Heart** | |
| --- | --- | --- | --- | --- | --- | --- | --- | --- | --- | --- | --- |
|  | **Deaths** | **MR^c^** |  | **Deaths** | **MR^c^** |  | **Deaths** | **MR^c^** |  | **Deaths** | **MR^c^** |
| Overall | 32,116 | 3,277 |  | 12,300 | 3,795 |  | 6,612 | 10,157 |  | 5,578 | 4,761 |
| Heart disease | 6,840 | 698 |  | 1,289 | 398 |  | 466 | 716 |  | 2,500 | 2,134 |
| Cancer | 4,697 | 479 |  | 3,166 | 977 |  | 1,033 | 1,586 |  | 867 | 740 |
| Kidney diseases | 4,451 | 454 |  | 632 | 195 |  | 191 | 293 |  | 297 | 253 |
| Diabetes mellitus | 3,772 | 385 |  | 335 | 103 |  | 42 | 65 |  | 141 | 121 |
| Other infections^d^ | 2,048 | 209 |  | 1,134 | 350 |  | 320 | 492 |  | 267 | 228 |
| Liver diseases | 519 | 53 |  | 2,684 | 828 |  | 45 | 69 |  | 37 | 31 |
| Other respiratory diseases | 670 | 68 |  | 255 | 79 |  | 1,882 | 2,891 |  | 131 | 112 |
| Cerebrovascular diseases | 1,255 | 128 |  | 317 | 98 |  | 96 | 148 |  | 150 | 128 |
| Other circulatory diseases | 1,310 | 134 |  | 201 | 62 |  | 61 | 93 |  | 90 | 76 |
| Influenza and pneumonia | 907 | 93 |  | 236 | 73 |  | 300 | 461 |  | 126 | 107 |
| Accidents and adverse events | 815 | 83 |  | 396 | 122 |  | 75 | 115 |  | 110 | 94 |
| Chronic lower respiratory diseases | 451 | 46 |  | 172 | 53 |  | 796 | 1,224 |  | 78 | 67 |
| Dementia and Alzheimer’s | 341 | 35 |  | 63 | 19 |  | 18 | 27 |  | 36 | 30 |
| Cystic fibrosis | 4 | <1 |  | 13 | 4 |  | 389 | 598 |  | 0 | 0 |
| Suicide and self-inflicted injury | 140 | 14 |  | 86 | 27 |  | 15 | 22 |  | 14 | 12 |
| Other cause of death | 3,897 | 398 |  | 1,322 | 408 |  | 883 | 1,357 |  | 735 | 627 |

MR, mortality rate; US, United States.

a. All recipients of a specified organ, regardless of whether other organs were also transplanted.
b. Since there is no International Classification of Diseases 10th Revision (ICD-10) code for graft failure, deaths classified as due to graft failure in the primary analyses were recategorized into the 16 cause of death groups accordingly to their NDI-assigned ICD-10 codes.
c. Deaths per 100,000 person-years.
d. This category includes all infections (viral, bacterial, fungal, and parasitic) except influenza and pneumonia which are classified as a separate cause of death group.

### eTable 22. SMRs for deaths among adult kidney and non-kidney recipients in the US, 1999–2019

| **Cause of death** | **Kidney** | | |  | **Recipients of organs other than kidney** | | |
| --- | --- | --- | --- | --- | --- | --- | --- |
|  | **Deaths** | **MR^a^** | **SMR (95% CI)** |  | **Deaths** | **MR^a^** | **SMR (95% CI)** |
| Overall | 85,434 | 3,511.9 | 3.46 (3.43–3.49) |  | 68,087 | 5,807.6 | 5.05 (5.00–5.10) |
| Heart disease | 18,252 | 750.3 | 3.14 (3.08–3.20) |  | 11,531 | 983.5 | 3.66 (3.58–3.76) |
| Cancer | 11,460 | 471.1 | 1.58 (1.54–1.62) |  | 12,458 | 1,062.6 | 2.95 (2.88–3.02) |
| Kidney diseases | 12,754 | 524.3 | 24.3 (23.7–24.9) |  | 2,394 | 204.2 | 9.92 (9.39–10.5) |
| Diabetes mellitus | 11,088 | 455.8 | 11.2 (11.0–11.5) |  | 1,218 | 103.9 | 2.41 (2.23–2.60) |
| Other infections | 5,695 | 234.1 | 6.72 (6.48–6.95) |  | 8,062 | 687.6 | 20.0 (19.4–20.5) |
| Liver diseases | 1,371 | 56.4 | 1.97 (1.84–2.11) |  | 9,093 | 775.6 | 21.3 (20.8–21.9) |
| Other respiratory diseases | 1,534 | 63.0 | 3.24 (3.03–3.46) |  | 5,795 | 494.3 | 21.8 (21.1–22.5) |
| Cerebrovascular diseases | 3,167 | 130.2 | 2.80 (2.67–2.94) |  | 1,441 | 122.9 | 2.67 (2.49–2.86) |
| Other circulatory diseases | 3,714 | 152.7 | 6.55 (6.26–6.85) |  | 905 | 77.2 | 3.21 (2.94–3.50) |
| Influenza and pneumonia | 2,167 | 89.1 | 5.38 (5.08–5.69) |  | 1,529 | 130.4 | 7.40 (6.93–7.90) |
| Accidents and adverse events | 2,055 | 84.5 | 1.61 (1.52–1.71) |  | 1,331 | 113.6 | 2.10 (1.95–2.25) |
| Chronic lower respiratory diseases | 1,067 | 43.9 | 0.83 (0.76–0.90) |  | 3,239 | 276.2 | 4.06 (3.88–4.25) |
| Dementia and Alzheimer's | 647 | 26.6 | 0.90 (0.81–1.00) |  | 199 | 17.0 | 0.62 (0.51–0.75) |
| Cystic fibrosis | 21 | 0.8 | 8.97 (4.86–16.6) |  | 1,308 | 111.6 | 1177 (1097–1262) |
| Suicide and self-inflicted injury | 432 | 17.7 | 1.07 (0.94–1.21) |  | 293 | 25.0 | 1.20 (1.04–1.39) |
| Other cause | 10,011 | 411.5 | 4.32 (4.21–4.44) |  | 7,291 | 621.9 | 5.84 (5.67–6.02) |

CI, confidence interval; MR, mortality rate; SMR, standardized mortality ratio; US, United States.

a. Deaths per 100,000 person-years.

### eTable 23. SMRs for deaths among adult liver and non-liver recipients in the US, 1999–2019

| **Cause of death** | **Liver** | | |  | **Recipients of organs other than liver** | | |
| --- | --- | --- | --- | --- | --- | --- | --- |
|  | **Deaths** | **MR^a^** | **SMR (95% CI)** |  | **Deaths** | **MR^a^** | **SMR (95% CI)** |
| Overall | 37,379 | 4,896.7 | 4.38 (4.32–4.45) |  | 116,142 | 4,087.0 | 3.91 (3.88–3.94) |
| Heart disease | 2,904 | 380.5 | 1.47 (1.40–1.55) |  | 26,879 | 945.9 | 3.85 (3.78–3.91) |
| Cancer | 8,483 | 1,111.3 | 3.19 (3.10–3.28) |  | 15,435 | 543.2 | 1.75 (1.71–1.78) |
| Kidney diseases | 1,479 | 193.8 | 9.74 (9.09–10.5) |  | 13,669 | 481.0 | 22.24 (21.7–22.8) |
| Diabetes mellitus | 787 | 103.1 | 2.42 (2.21–2.67) |  | 11,519 | 405.4 | 9.87 (9.62–10.1) |
| Other infections | 6,699 | 877.6 | 25.7 (24.8–26.5) |  | 7,058 | 248.4 | 7.12 (6.90–7.35) |
| Liver diseases | 9,451 | 1,238.1 | 32.9 (32.1–33.8) |  | 1,013 | 35.7 | 1.21 (1.12–1.32) |
| Other respiratory diseases | 595 | 77.9 | 3.58 (3.22–3.97) |  | 6,734 | 237.0 | 11.7 (11.4–12.1) |
| Cerebrovascular diseases | 817 | 107.0 | 2.38 (2.17–2.60) |  | 3,792 | 133.4 | 2.86 (2.74–2.99) |
| Other circulatory diseases | 541 | 70.9 | 3.05 (2.73–3.40) |  | 4,078 | 143.5 | 6.07 (5.82–6.34) |
| Influenza and pneumonia | 500 | 65.5 | 3.81 (3.40–4.27) |  | 3,196 | 112.5 | 6.68 (6.38–7.00) |
| Accidents and adverse events | 947 | 124.1 | 2.33 (2.14–2.53) |  | 2,439 | 85.8 | 1.62 (1.54–1.71) |
| Chronic lower respiratory diseases | 376 | 49.3 | 0.77 (0.67–0.88) |  | 3,929 | 138.3 | 2.46 (2.36–2.57) |
| Dementia and Alzheimer's | 104 | 13.7 | 0.51 (0.39–0.67) |  | 741 | 26.1 | 0.89 (0.81–0.98) |
| Cystic fibrosis | 26 | 3.4 | 39.3 (24.0–64.3) |  | 1,303 | 45.8 | 473 (441–508) |
| Suicide and self-inflicted injury | 222 | 29.1 | 1.41 (1.20–1.67) |  | 503 | 17.7 | 1.02 (0.91–1.15) |
| All other codes | 3,447 | 451.5 | 4.35 (4.16–4.54) |  | 13,855 | 487.6 | 5.00 (4.89–5.11) |

CI, confidence interval; MR, mortality rate; SMR, standardized mortality ratio; US, United States.

a. Deaths per 100,000 person-years.

### eTable 24. SMRs for deaths among adult lung and non-lung recipients in the US, 1999–2019

| **Cause of death** | **Lung** | | |  | **Recipients of organs other than lung** | | |
| --- | --- | --- | --- | --- | --- | --- | --- |
|  | **Deaths** | **MR^a^** | **SMR (95% CI)** |  | **Deaths** | **MR^a^** | **SMR (95% CI)** |
| Overall | 17,379 | 11,493.1 | 10.43 (10.2–10.6) |  | 136,142 | 3,941.7 | 3.73 (3.70–3.75) |
| Heart disease | 1,118 | 739.7 | 2.98 (2.77–3.21) |  | 28,665 | 829.9 | 3.34 (3.29–3.39) |
| Cancer | 2,184 | 1,444.0 | 4.02 (3.82–4.24) |  | 21,735 | 629.3 | 1.98 (1.95–2.02) |
| Kidney diseases | 386 | 255.3 | 13.43 (11.9–15.2) |  | 14,762 | 427.4 | 20.0 (19.6–20.5) |
| Diabetes mellitus | 80 | 52.8 | 1.32 (1.00–1.74) |  | 12,226 | 354.0 | 8.54 (8.34–8.75) |
| Other infections | 901 | 595.7 | 19.12 (17.6–20.8) |  | 12,856 | 372.2 | 10.7 (10.4–0.92) |
| Liver diseases | 125 | 82.4 | 2.54 (2.03–3.17) |  | 10,340 | 299.4 | 9.64 (9.40–9.89) |
| Other respiratory diseases | 4,976 | 3,291.1 | 150 (144–155) |  | 2,352 | 68.1 | 3.33 (3.16–3.51) |
| Cerebrovascular diseases | 244 | 161.3 | 3.85 (3.29–4.50) |  | 4,364 | 126.4 | 2.72 (2.61–2.83) |
| Other circulatory diseases | 143 | 94.7 | 4.25 (3.47–5.21) |  | 4,476 | 129.6 | 5.49 (5.27–5.72) |
| Influenza and pneumonia | 802 | 530.3 | 32.4 (29.7–35.4) |  | 2,894 | 83.8 | 4.95 (4.71–5.20) |
| Accidents and adverse events | 153 | 101.0 | 2.00 (1.63–2.45) |  | 3,234 | 93.6 | 1.76 (1.68–1.84) |
| Chronic lower respiratory diseases | 2,744 | 1,814.8 | 25.5 (24.2–26.8) |  | 1,562 | 45.2 | 0.79 (0.74–0.85) |
| Dementia and Alzheimer's | 22 | 14.3 | 0.62 (0.38–1.02) |  | 824 | 23.9 | 0.82 (0.75–0.90) |
| Cystic fibrosis | 1,298 | 858.6 | 7420 (6920–7955) |  | 30 | 0.9 | 9.44 (5.72–15.6) |
| Suicide and self-inflicted injury | 34 | 22.7 | 1.15 (0.76–1.76) |  | 690 | 20.0 | 1.11 (1.01–1.23) |
| All other codes | 2,169 | 1,434.6 | 13.92 (13.2–14.7) |  | 15,133 | 438.1 | 4.44 (4.34–4.54) |

CI, confidence interval; MR, mortality rate; SMR, standardized mortality ratio; US, United States.

a. Deaths per 100,000 person-years.

### eTable 25. SMRs for deaths among adult heart and non-heart recipients in the US, 1999–2019

| **Cause of death** | **Heart** | | |  | **Recipients of organs other than heart** | | |
| --- | --- | --- | --- | --- | --- | --- | --- |
|  | **Deaths** | **MR^a^** | **SMR (95% CI)** |  | **Deaths** | **MR^a^** | **SMR (95% CI)** |
| Overall | 15,626 | 5,451.0 | 4.16 (4.07–4.25) |  | 137,895 | 4,155.5 | 4.00 (3.97–4.03) |
| Heart disease | 7,970 | 2,780.2 | 8.77 (8.51–9.04) |  | 21,813 | 657.3 | 2.71 (2.66–2.76) |
| Cancer | 2,138 | 745.7 | 1.83 (1.73–1.94) |  | 21,780 | 656.4 | 2.11 (2.07–2.15) |
| Kidney diseases | 696 | 242.8 | 9.89 (8.93–11.0) |  | 14,452 | 435.5 | 20.8 (20.3–21.2) |
| Diabetes mellitus | 363 | 126.7 | 2.59 (2.25–2.98) |  | 11,942 | 359.9 | 8.84 (8.62–9.06) |
| Other infections | 799 | 278.7 | 7.20 (6.57–7.90) |  | 12,958 | 390.5 | 11.4 (11.1–11.6) |
| Liver diseases | 93 | 32.5 | 0.89 (0.69–1.16) |  | 10,371 | 312.5 | 10.2 (9.95–10.5) |
| Other respiratory diseases | 305 | 106.5 | 4.03 (3.47–4.66) |  | 7,024 | 211.7 | 10.6 (10.3–11.0) |
| Cerebrovascular diseases | 409 | 142.7 | 2.67 (2.34–3.05) |  | 4,199 | 126.5 | 2.77 (2.66–2.89) |
| Other circulatory diseases | 270 | 94.2 | 3.34 (2.83–3.94) |  | 4,349 | 131.1 | 5.66 (5.44–5.90) |
| Influenza and pneumonia | 268 | 93.4 | 4.64 (3.95–5.46) |  | 3,428 | 103.3 | 6.21 (5.94–6.49) |
| Accidents and adverse events | 265 | 92.3 | 1.56 (1.33–1.84) |  | 3,122 | 94.1 | 1.79 (1.71–1.88) |
| Chronic lower respiratory diseases | 144 | 50.2 | 0.64 (0.51–0.80) |  | 4,162 | 125.4 | 2.23 (2.14–2.33) |
| Dementia and Alzheimer's | 70 | 24.5 | 0.77 (0.56–1.07) |  | 776 | 23.4 | 0.82 (0.75–0.90) |
| Cystic fibrosis | 5 | 1.7 | 18.7 (5.43–62.1) |  | 1,324 | 39.9 | 420 (392–451) |
| Suicide and self-inflicted injury | 42 | 14.8 | 0.68 (0.46–1.00) |  | 682 | 20.6 | 1.16 (1.05–1.28) |
| All other codes | 1,789 | 624.0 | 5.26 (4.95–5.58) |  | 15,513 | 467.5 | 4.81 (4.71–4.91) |

CI, confidence interval; MR, mortality rate; SMR, standardized mortality ratio; US, United States.

a. Deaths per 100,000 person-years.
